# Precision medicine in type 2 diabetes: A systematic review of treatment effect heterogeneity for GLP1-receptor agonists and SGLT2-inhibitors

**DOI:** 10.1101/2023.04.21.23288868

**Authors:** Katherine G Young, Eram Haider McInnes, Robert J Massey, Anna R Kahkohska, Scott J Pilla, Sridharan Raghaven, Maggie A Stanislawski, Deirdre K Tobias, Andrew P McGovern, Adem Y Dawed, Angus G Jones, Ewan R Pearson, John M Dennis, ADA/EASD Precision Medicine in Diabetes Initiative Consortium

## Abstract

**Background:** A precision medicine approach in type 2 diabetes requires identification of clinical and biological features that are reproducibly associated with differences in clinical outcomes with specific anti-hyperglycaemic therapies. Robust evidence of such treatment effect heterogeneity could support more individualized clinical decisions on optimal type 2 diabetes therapy.

**Methods:** We performed a pre-registered systematic review of meta-analysis studies, randomized control trials, and observational studies evaluating clinical and biological features associated with heterogenous treatment effects for SGLT2-inhibitor and GLP1-receptor agonist therapies, considering glycaemic, cardiovascular, and renal outcomes.

**Results:** After screening 5,686 studies, we included 101 studies of SGLT2-inhibitors and 75 studies of GLP1-receptor agonists in the final systematic review. The majority of papers had methodological limitations precluding robust assessment of treatment effect heterogeneity. For glycaemic outcomes, most cohorts were observational, with multiple analyses identifying lower renal function as a predictor of lesser glycaemic response with SGLT2-inhibitors and markers of reduced insulin secretion as predictors of lesser response with GLP1-receptor agonists. For cardiovascular and renal outcomes, the majority of included studies were post-hoc analyses of randomized control trials (including meta-analysis studies) which identified limited clinically relevant treatment effect heterogeneity.

**Conclusions:** Current evidence on treatment effect heterogeneity for SGLT2-inhibitor and GLP1-receptor agonist therapies is limited, likely reflecting the methodological limitations of published studies. Robust and appropriately powered studies are required to understand type 2 diabetes treatment effect heterogeneity and evaluate the potential for precision medicine to inform future clinical care.

**Plain language summary:** This review identifies research that helps understand which clinical and biological factors that are associated with different outcomes for specific type 2 diabetes treatments. This information could help clinical providers and patients make better informed personalized decisions about type 2 diabetes treatments. We focused on two common type 2 diabetes treatments: SGLT2-inhibitors and GLP1-receptor agonists, and three outcomes: blood glucose control, heart disease, and kidney disease. We identified some potential factors that are likely to lessen blood glucose control including lower kidney function for SGLT2-inhibitors and lower insulin secretion for GLP1-receptor agonists. We did not identify clear factors that alter heart and renal disease outcomes for either treatment. Most of the studies had limitations, meaning more research is needed to fully understand the factors that influence treatment outcomes in type 2 diabetes.

## INTRODUCTION

Two of the most recently introduced anti-hyperglycaemic drug classes, SGLT2-inhibitors (SGLT2i) and GLP1-receptor agonists (GLP1-RA), have been shown in randomized clinical trials not only to reduce glycaemia^1^ but also to lower the risk of renal and cardiovascular disease (CVD) outcomes among high-risk individuals with type 2 diabetes (T2D)^2–5^. Based on average treatment effects reported in placebo-controlled trials, current T2D clinical consensus guidelines recommend a stratified approach to treatment selection, preferentially recommending these drug classes independent of their glucose lowering effect for individuals with cardiovascular or renal comorbidity. Specifically, people with heart failure and/or chronic kidney disease are recommended to initiate SGLT2i and people with prior CVD or high risk for CVD are recommended to initiate either an SGLT2i or a GLP1-RA. In addition, these drugs are recommended as second-line glucose lowering medications to be added after metformin^6^.

A limitation of the current stratified approach to SGLT2i and GLP1-RA treatment in clinical guidelines is that is informed by trial recruitment strategies, and consequential accumulation of evidence of treatment benefits only for specific subgroups with or at high risk of cardiorenal disease, rather than from an understanding of how the benefits and risks of each drug class vary across the whole spectrum of T2D. A more comprehensive approach to treatment selection would require recognition of the extreme heterogeneity in the demographic, clinical, and biological features of people with T2D, and the impact of this heterogeneity on drug-specific clinical outcomes. Identification of robust and reproducible patterns of heterogenous treatment effects is plausible as, at the individual patient level, responses to the same drug treatment appears to vary greatly^7^. A greater understanding of population-wide heterogenous treatment effects and enhanced capacity to predict individual treatment responses is needed to advance towards the central goal of precision type 2 diabetes medicine—where demographic, clinical, biological, or other patient-level features may be used to match individuals to their optimal anti-hyperglycaemic regimen as part of routine T2D diabetes care.

To assess the evidence base for treatment effect heterogeneity for SGLT2i and GLP1-RA, we undertook a systematic literature review to summarize key findings from studies that specifically examined interactions between individual-level biomarkers and the effects of these drug classes on clinical outcomes. Although biomarkers may connote laboratory-based measurements in traditional contexts, herein we broadly conceptualized biomarkers as individual-level demographic, clinical, and biological features including both laboratory measures as well as genetic and genomic markers. We focused on three categories of outcomes relevant to T2D care: (1) glycaemic response (as measured by hemoglobin A1c; HbA1c); (2) CVD outcomes; and (3) renal outcomes. Our review was guided by the following research question: In a population with T2D, treated with SGLT2i or GLP1-RA, what are the biomarkers associated with heterogenous treatment effects in glycaemic, CVD, and renal outcomes? Each of the three outcomes were evaluated separately for each of the two drug classes for a total of 6 sub-studies. Given the heterogeneity of the T2D population, we anticipated that we would find one or more biomarkers modifying the effects of SGLT2i and GLP1-RA.

The Precision Medicine in Diabetes Initiative (PMDI) was established in 2018 by the American Diabetes Association (ADA) in partnership with the European Association for the Study of Diabetes (EASD). The ADA/EASD PMDI includes global thought leaders in precision diabetes medicine who are working to address the burgeoning need for higher quality, individualized diabetes prevention and care through precision medicine^8^. This Systematic Review is written on behalf of the ADA/EASD PMDI as part of a comprehensive evidence evaluation in support of the 2nd International Consensus Report on Precision Diabetes Medicine.

## METHODS

We conducted a systematic review according to the Preferred Reporting Items for Systematic Reviews and Meta-Analyses (PRISMA) guidelines^9^. See Appendix 1 for a checklist of each component. We first developed and iterated a protocol for the review (CRD42022303236). As above, our review was guided by the following research question: In a population with T2D, treated with SGLT2i and GLP1-RA, what are the biomarkers associated with heterogenous treatment effects in glycaemic, CVD, and renal outcomes?

### Search Strategy

The search strategy for this review was developed for each drug class (SGLT2i and GLP1-RA) and outcome (glycaemic, cardiovascular, and renal) to capture studies specifically evaluating treatment effect heterogeneity associated with demographic, clinical, and biological features in people with type 2 diabetes. Potential effect modifiers of interest comprised age, sex, ethnicity, clinical features, routine blood tests, metabolic markers, and genetics; all search terms were based on medical subject sub-headings (MeSH) terms and are reported in **Supplementary Material section 1**. SGLT2i and GLP1-RA were evaluated at drug class level and we did not aim to identify within-class heterogeneity in treatment effects. Electronic searches were performed in PubMed and Embase by two independent academic librarians in February 2022. Forwards and backwards citation searching was conducted but grey literature and white papers were not searched.

### Inclusion criteria

To be included, studies were required to meet the following criteria:

1. Full-text English-language publications of RCTs, meta-analyses, post-hoc analyses of RCTs, pooled cohort analyses, prospective and retrospective observational analyses published in peer-reviewed journals.
2. Adult populations with type 2 diabetes taking at least one of either SGLT2i or GLP1-RA with sample size >100 for the active drug of interest.
3. At least a 4 month potential follow up period after initiation of the drug class of interest.
4. Randomized control trials (RCTs) required a comparison against placebo or an active comparator antihyperglycaemic drug. Observational studies did not require a comparator group.
5. Pre-specified aim of the study to examine heterogeneity in treatment outcome, such as biomarker-treatment interactions, stratified analyses, or heterogeneity-focused machine learning approaches.
6. Reported differential effects of the drug class on an outcome of interest with respect to a biomarker.
7. All individual trial or observational cohorts included in a meta-analysis or pooled cohort analysis must have met the inclusion criteria stated above.

We further excluded studies based on the following criteria: studied type 1 or other forms of non-type 2 diabetes; included children/minors; inpatient studies; conference proceeding abstracts, editorials, opinions papers, book chapters, clinical trial registries, case reports, commentaries, narrative reviews, or non-peer reviewed studies; did not adequately adjust for confounders (individual RCTs and observational studies only, this criteria was not applied for meta-analyses and pooled cohort analyses); did not address the question of treatment response heterogeneity for biomarkers of interest.

Titles and abstracts were independently screened by pairs of research team members to identify potentially eligible studies; these were then independently evaluated for inclusion in the full-text review. Any discrepancies were discussed with a third author until reaching consensus. Discrepancies were discussed as part of larger group meetings to ensure consistency in decisions across reviewer pairs.

### Data Extraction and Quality Assessment

Pairs of authors independently reviewed the main reports and supplementary materials and extracted the following data for each of the included papers:

1. Publication (PMID, journal, publication year, first author, title, study type)
2. Study (setting and region, study time period, follow up period)
3. Population (overall characteristics, ethnicity)
4. Intervention (Drug class, specific therapies, treatment/comparator arm sizes)
5. Statistical analysis (outcome, outcome measurement, subgroups/predictors analysed with respect to biomarkers, statistical model, covariate set)
6. Results (relevant figures and tables, main findings, methodology, quality)

After data were extracted, information was synthesized by drug class and outcome and further examined by biomarkers or subgroups analyzed within each study. Results were extracted within these subsections and summarized for each paper, where general trends in results for each subsection were outlined.

Risk of bias evaluations using the Joanna Briggs Institute (JBI) critical appraisal tool for cohort studies^10^ was also conducted alongside the data extraction by each pair of authors. This was used to determine the extent of bias within the study’s design, execution, and analysis, specifically within the population, outcome measurements, and statistical modelling. Further detail on the risk of bias can be seen in **Supplementary Figure 2**. Additionally, the Grading of Recommendations, Assessment, Development, and Evaluations (GRADE) framework^11^ was applied at the outcome level for each drug class to determine the quality of evidence and certainty of effects for these subsections; an overall GRADE evaluation for all evidence was also provided.

### Outcomes

Three outcome categories were assessed in the included studies: (1) changes in HbA1c associated with treatment; (2) CVD outcomes limited to cardiovascular (CV)-related death, non-fatal myocardial infarction, non-fatal stroke, hospitalization for angina, coronary artery bypass graft, percutaneous coronary intervention, hospitalization for heart failure, carotid endarterectomy, and peripheral vascular disease; and (3) renal outcomes including development of chronic kidney disease, and longitudinal changes in markers of renal function including eGFR and albuminuria. Specific measurements and their procedures for each category of outcome varied across the included studies. Summaries of the included papers assessing each outcome for each drug class are reported in **Supplementary Tables 1a-f**.

## RESULTS

### Literature search and screening results

**Supplementary Figure 1** depicts the outcomes of the study screening processes for SGLT2i (**1a**) and GLP1-RA (**1b**).

For SGLT2i, a total of 3415 unique citations underwent title and abstract screening. 3076 were determined to not meet the pre-defined eligibility criteria. The remaining 339 full-text articles were screened, through which process 238 articles were excluded. The most common reasons for exclusion included studies which did not report on heterogeneity of treatment response (126 studies), studies reporting only univariate or unadjusted associations (41 studies), and studies that did not meet inclusion criteria (64 studies). In total, 101 studies were identified for inclusion based on the systematic search.

For GLP1-RA, a total of 2270 unique citations underwent title and abstract screening. 2109 were determined to not meet the pre-defined eligibility criteria. The remaining 161 full-text articles were screened, through which process 86 articles were excluded. The most common reasons for exclusion included studies that did not meet inclusion criteria (39 studies), and studies reporting only univariate or unadjusted associations (26 studies), and studies which did not report on heterogeneity of treatment response (17 studies). In total, 75 studies were identified for inclusion.

### Description of included studies

Included studies for CVD and renal outcomes were predominantly secondary analyses of industry-funded placebo-controlled trials (RCT), or meta-analyses of these trials, with a smaller number of observational studies. For glycaemic outcomes, most studies were observational. **Supplementary Table 1 (a-f)** shows all included studies – for each of GLP1-RA and SGLT2i, split by glycaemic, CVD, and renal outcomes, and including information on study population size, examined biomarkers, and significant findings. Summaries of the individual RCTs informing included meta-analyses are detailed in **Supplementary Table 2**.

### SGLT2i, GLP1-RA AND GLYCAEMIC OUTCOMES

Study quality for assessment of heterologous treatment effects for both SGLT2i and GLP1-RA was variable with strong methodological limitations for the study of predictors of glycaemia treatment response common. A core weakness with many studies was a lack of head-to-head comparisons between therapies, which is required to separate broader prognostic factors (that predict response to any glucose-lowering therapy) from drug-specific factors that are associated with differential treatment response. Put otherwise, even when data suggested that a biomarker was associated with glycaemic response, it was not clear if this factor was helpful for choosing between therapies due to the lack of an active comparator. Other common methodological weaknesses included the use of arbitrary subgroups (rather than assessment of continuous predictors), small numbers in comparator subgroups which limited statistical power, dichotomized outcomes (responder analysis), multiple testing, and lack of adjustment for key potential confounders.

#### SGLT2i

Of 27 studies that met our inclusion/exclusion criteria, 9 observational studies (usually retrospective analysis of healthcare records), 5 post-hoc analysis of individual randomized control trials (RCTs), 10 pooled analyses of individual data from multiple RCTs, and 3 RCT meta-analyses were included (**Supplementary Table 1e**). All included studies assessed routine clinical characteristics and routinely measured clinical biomarkers (**Table 1**). No pharmacogenetic, or, with the exception of one study of HOMA-B,^12^ non-routine biomarker studies were identified.

**Table 1:**
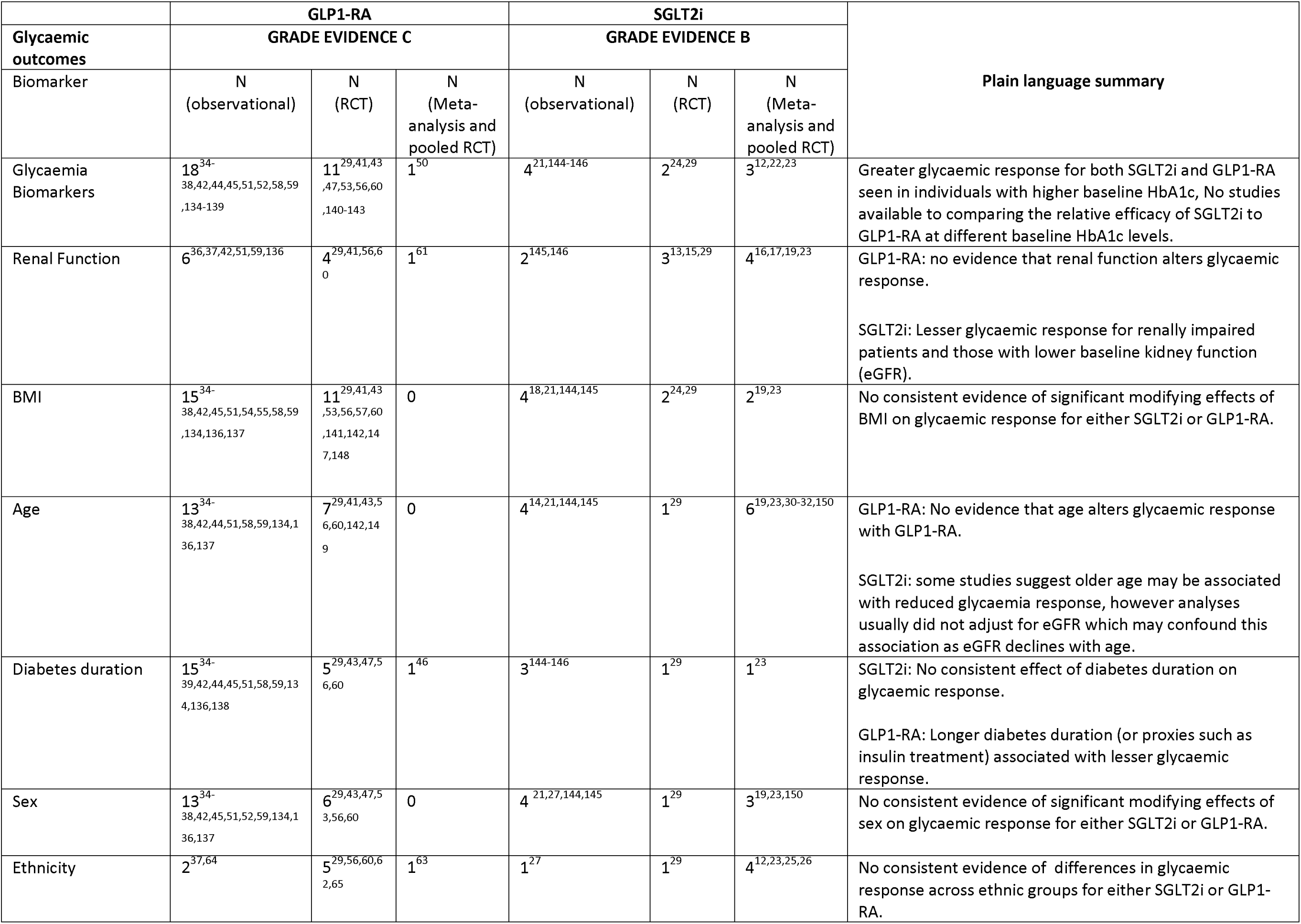
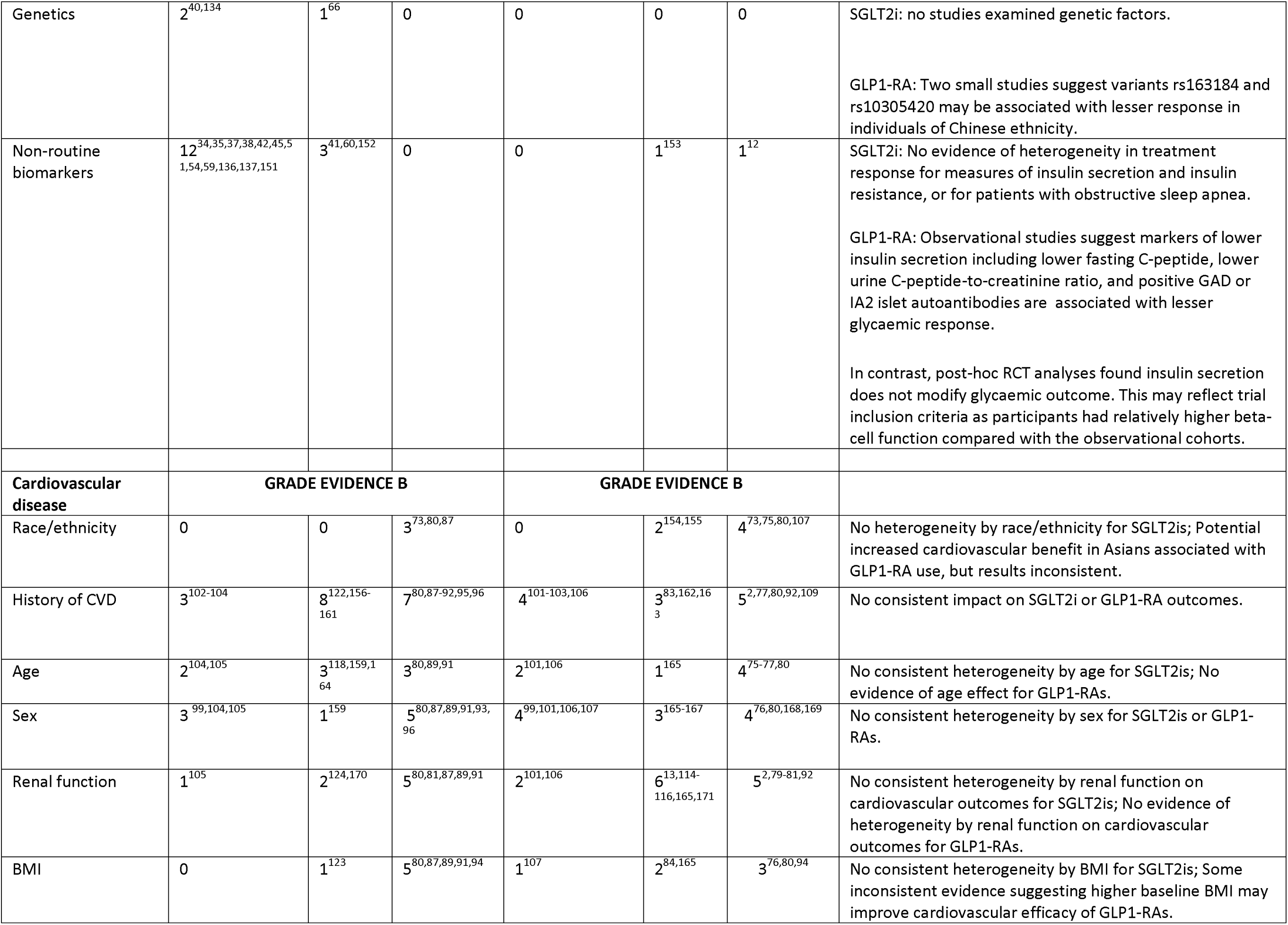
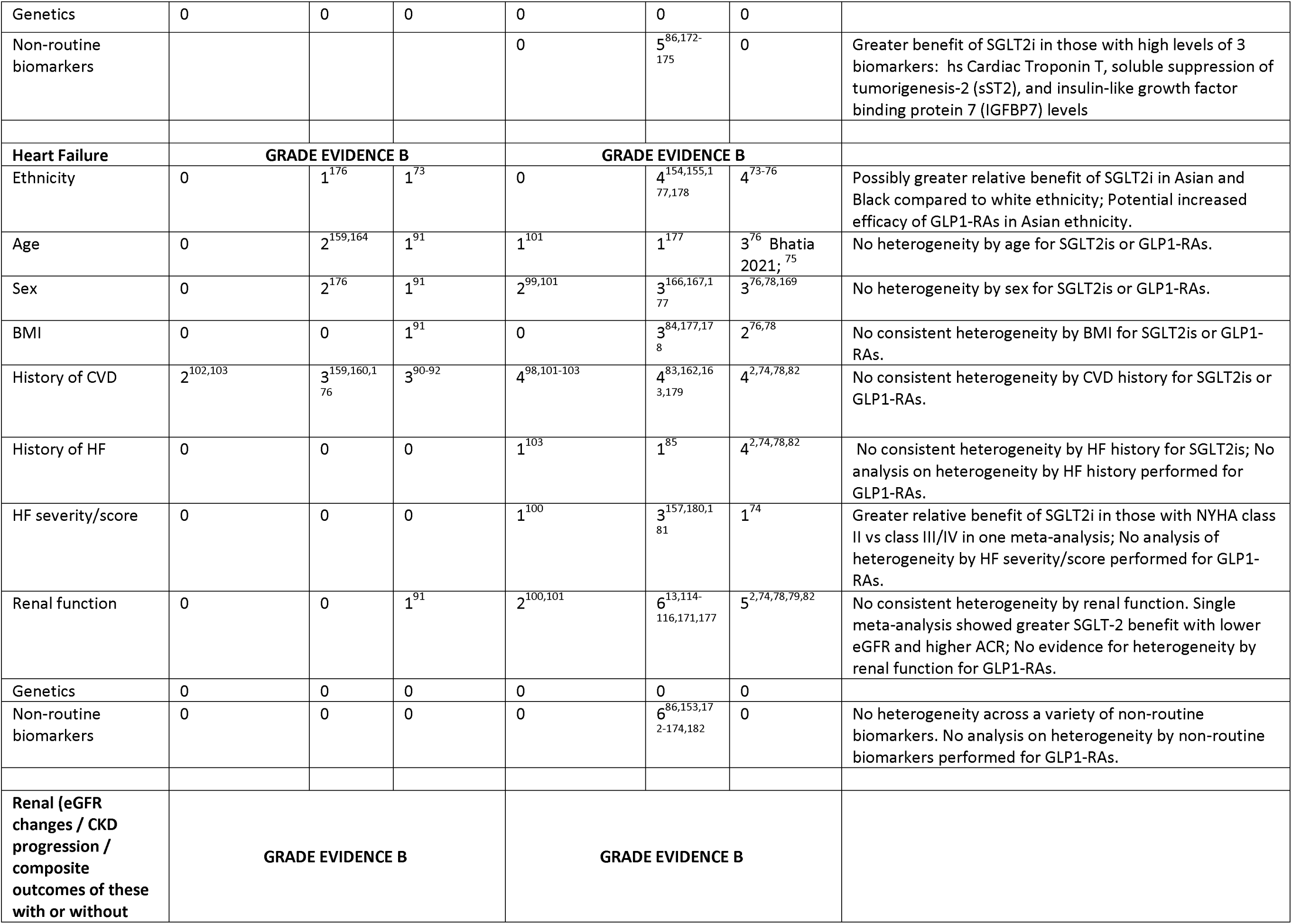
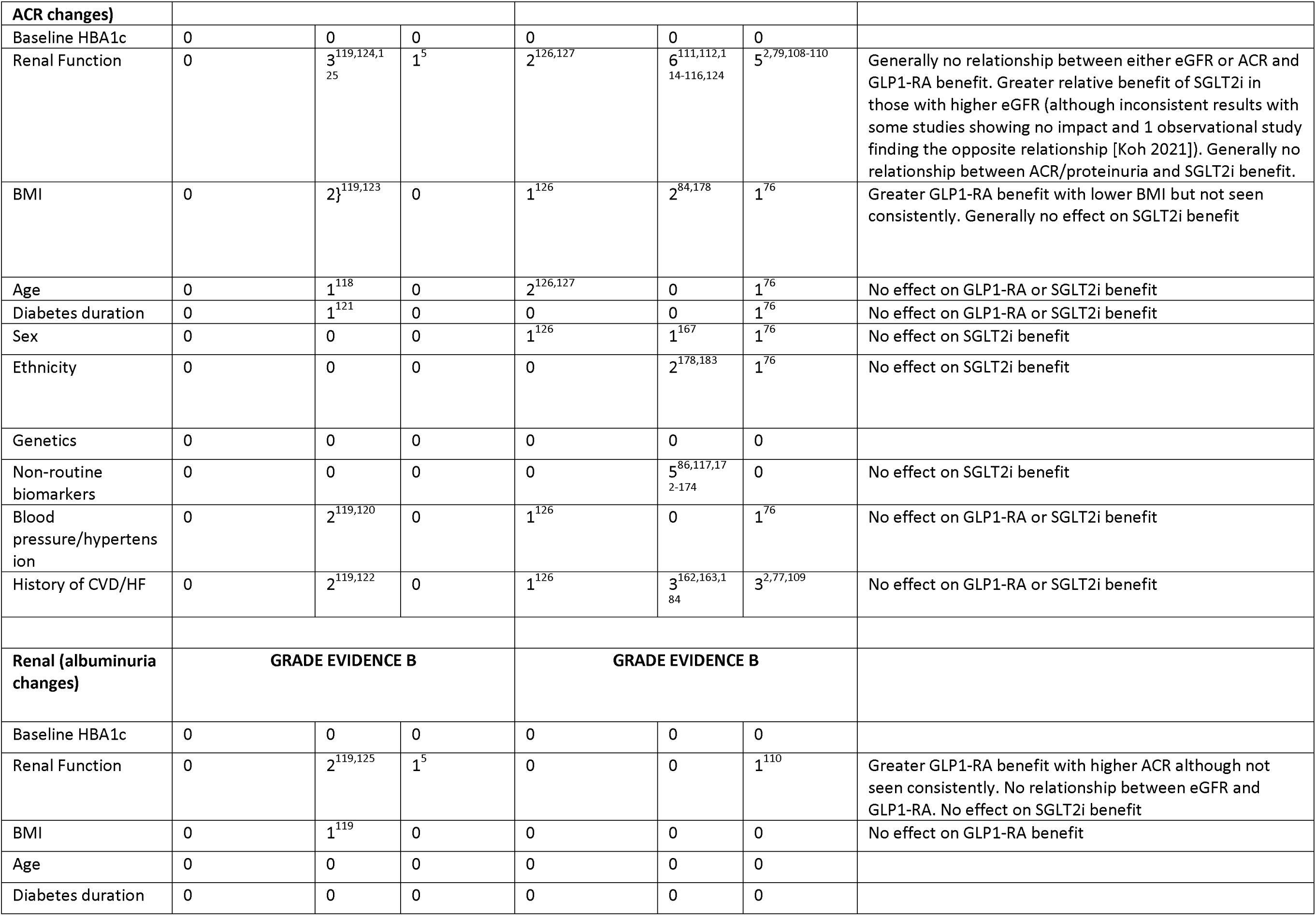
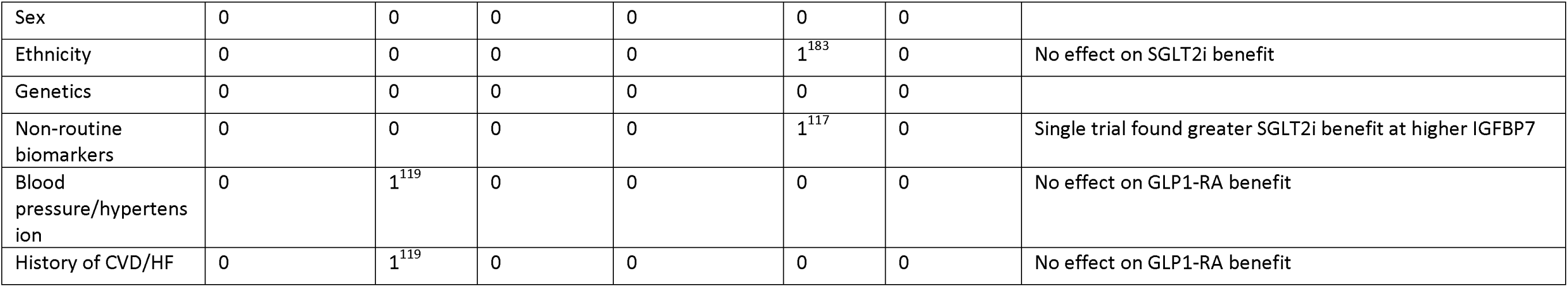
Summary of evidence for treatment effect heterogeneity for SGLT2-inhibitor and GLP1-receptor agonist therapies for glycaemic, cardiovascular (including heart failure), and renal outcomes

|  | GLP1-RA |  |  | SGLT2i |  |  | Plain language summary |
| --- | --- | --- | --- | --- | --- | --- | --- |
| Glycaemic outcomes | GRADE EVIDENCE C |  |  | GRADE EVIDENCE B |  |  |  |
| Biomarker | N (observational) | N (RCT) | N (Meta-analysis and pooled RCT) | N (observational) | N (RCT) | N (Meta-analysis and pooled RCT) |  |
| Glycaemia Biomarkers | 18<sup>34-38,42,44,45,51,52,58,59,134-139</sup> | 11<sup>29,41,43,47,53,56,60,140-143</sup> | 1<sup>50</sup> | 4<sup>21,144-146</sup> | 2<sup>24,29</sup> | 3<sup>12,22,23</sup> | Greater glycaemic response for both SGLT2i and GLP1-RA seen in individuals with higher baseline HbA1c, No studies available to comparing the relative efficacy of SGLT2i to GLP1-RA at different baseline HbA1c levels. |
| Renal Function | 6<sup>36,37,42,51,59,136</sup> | 4<sup>29,41,56,60</sup> | 1<sup>61</sup> | 2<sup>145,146</sup> | 3<sup>13,15,29</sup> | 4<sup>16,17,19,23</sup> | GLP1-RA: no evidence that renal function alters glycaemic response.<br>SGLT2i: Lesser glycaemic response for renally impaired patients and those with lower baseline kidney function (eGFR). |
| BMI | 15<sup>34-38,42,45,51,54,55,58,59,134,136,137</sup> | 11<sup>29,41,43,53,56,57,60,141,142,147,148</sup> | 0 | 4<sup>18,21,144,145</sup> | 2<sup>24,29</sup> | 2<sup>19,23</sup> | No consistent evidence of significant modifying effects of BMI on glycaemic response for either SGLT2i or GLP1-RA. |
| Age | 13<sup>34-38,42,44,51,58,59,134,136,137</sup> | 7<sup>29,41,43,56,60,142,149</sup> | 0 | 4<sup>14,21,144,145</sup> | 1<sup>29</sup> | 6<sup>19,23,30-32,150</sup> | GLP1-RA: No evidence that age alters glycaemic response with GLP1-RA.<br>SGLT2i: some studies suggest older age may be associated with reduced glycaemia response, however analyses usually did not adjust for eGFR which may confound this association as eGFR declines with age. |
| Diabetes duration | 15<sup>34-39,42,44,45,51,58,59,134,136,138</sup> | 5<sup>29,43,47,56,60</sup> | 1<sup>46</sup> | 3<sup>144-146</sup> | 1<sup>29</sup> | 1<sup>23</sup> | SGLT2i: No consistent effect of diabetes duration on glycaemic response.<br>GLP1-RA: Longer diabetes duration (or proxies such as insulin treatment) associated with lesser glycaemic response. |
| Sex | 13<sup>34-38,42,45,51,52,59,134,136,137</sup> | 6<sup>29,43,47,53,56,60</sup> | 0 | 4<sup>21,27,144,145</sup> | 1<sup>29</sup> | 3<sup>19,23,150</sup> | No consistent evidence of significant modifying effects of sex on glycaemic response for either SGLT2i or GLP1-RA. |
| Ethnicity | 2<sup>37,64</sup> | 5<sup>29,56,60,62,65</sup> | 1<sup>63</sup> | 1<sup>27</sup> | 1<sup>29</sup> | 4<sup>12,23,25,26</sup> | No consistent evidence of differences in glycaemic response across ethnic groups for either SGLT2i or GLP1-RA. |
| Genetics | 2<sup>40,134</sup> | 1<sup>66</sup> | 0 | 0 | 0 | 0 | SGLT2i: no studies examined genetic factors.<br>GLP1-RA: Two small studies suggest variants rs163184 and rs10305420 may be associated with lesser response in individuals of Chinese ethnicity. |
| Non-routine biomarkers | 12<sup>34,35,37,38,42,45,51,54,59,136,137,151</sup> | 3<sup>41,60,152</sup> | 0 | 0 | 1<sup>153</sup> | 1<sup>12</sup> | SGLT2i: No evidence of heterogeneity in treatment response for measures of insulin secretion and insulin resistance, or for patients with obstructive sleep apnea.<br>GLP1-RA: Observational studies suggest markers of lower insulin secretion including lower fasting C-peptide, lower urine C-peptide-to-creatinine ratio, and positive GAD or IA2 islet autoantibodies are associated with lesser glycaemic response.<br>In contrast, post-hoc RCT analyses found insulin secretion does not modify glycaemic outcome. This may reflect trial inclusion criteria as participants had relatively higher beta-cell function compared with the observational cohorts. |
| **Cardiovascular disease** | **GRADE EVIDENCE B** |  |  | **GRADE EVIDENCE B** |  |  |  |
| Race/ethnicity | 0 | 0 | 3<sup>73,80,87</sup> | 0 | 2<sup>154,155</sup> | 4<sup>73,75,80,107</sup> | No heterogeneity by race/ethnicity for SGLT2is; Potential increased cardiovascular benefit in Asians associated with GLP1-RA use, but results inconsistent. |
| History of CVD | 3<sup>102-104</sup> | 8<sup>122,156-161</sup> | 7<sup>80,87-92,95,96</sup> | 4<sup>101-103,106</sup> | 3<sup>83,162,163</sup> | 5<sup>2,77,80,92,109</sup> | No consistent impact on SGLT2i or GLP1-RA outcomes. |
| Age | 2<sup>104,105</sup> | 3<sup>118,159,164</sup> | 3<sup>80,89,91</sup> | 2<sup>101,106</sup> | 1<sup>165</sup> | 4<sup>75-77,80</sup> | No consistent heterogeneity by age for SGLT2is; No evidence of age effect for GLP1-RAs. |
| Sex | 3<sup>99,104,105</sup> | 1<sup>159</sup> | 5<sup>80,87,89,91,93,96</sup> | 4<sup>99,101,106,107</sup> | 3<sup>165-167</sup> | 4<sup>76,80,168,169</sup> | No consistent heterogeneity by sex for SGLT2is or GLP1-RAs. |
| Renal function | 1<sup>105</sup> | 2<sup>124,170</sup> | 5<sup>80,81,87,89,91</sup> | 2<sup>101,106</sup> | 6<sup>13,114-116,165,171</sup> | 5<sup>2,79-81,92</sup> | No consistent heterogeneity by renal function on cardiovascular outcomes for SGLT2is; No evidence of heterogeneity by renal function on cardiovascular outcomes for GLP1-RAs. |
| BMI | 0 | 1<sup>123</sup> | 5<sup>80,87,89,91,94</sup> | 1<sup>107</sup> | 2<sup>84,165</sup> | 3<sup>76,80,94</sup> | No consistent heterogeneity by BMI for SGLT2is; Some inconsistent evidence suggesting higher baseline BMI may improve cardiovascular efficacy of GLP1-RAs. |
| Genetics | 0 | 0 | 0 | 0 | 0 | 0 |  |
| Non-routine biomarkers |  |  |  | 0 | 5<sup>86,172-175</sup> | 0 | Greater benefit of SGLT2i in those with high levels of 3 biomarkers: hs Cardiac Troponin T, soluble suppression of tumorigenesis-2 (sST2), and insulin-like growth factor binding protein 7 (IGFBP7) levels |
| **Heart Failure** | **GRADE EVIDENCE B** |  |  | **GRADE EVIDENCE B** |  |  |  |
| Ethnicity | 0 | 1<sup>176</sup> | 1<sup>73</sup> | 0 | 4<sup>154,155,177,178</sup> | 4<sup>73-76</sup> | Possibly greater relative benefit of SGLT2i in Asian and Black compared to white ethnicity; Potential increased efficacy of GLP1-RAs in Asian ethnicity. |
| Age | 0 | 2<sup>159,164</sup> | 1<sup>91</sup> | 1<sup>101</sup> | 1<sup>177</sup> | 3<sup>76</sup> Bhatia 2021;<sup>75</sup> | No heterogeneity by age for SGLT2is or GLP1-RAs. |
| Sex | 0 | 2<sup>176</sup> | 1<sup>91</sup> | 2<sup>99,101</sup> | 3<sup>166,167,177</sup> | 3<sup>76,78,169</sup> | No heterogeneity by sex for SGLT2is or GLP1-RAs. |
| BMI | 0 | 0 | 1<sup>91</sup> | 0 | 3<sup>84,177,178</sup> | 2<sup>76,78</sup> | No consistent heterogeneity by BMI for SGLT2is or GLP1-RAs. |
| History of CVD | 2<sup>102,103</sup> | 3<sup>159,160,176</sup> | 3<sup>90-92</sup> | 4<sup>98,101-103</sup> | 4<sup>83,162,163,179</sup> | 4<sup>2,74,78,82</sup> | No consistent heterogeneity by CVD history for SGLT2is or GLP1-RAs. |
| History of HF | 0 | 0 | 0 | 1<sup>103</sup> | 1<sup>85</sup> | 4<sup>2,74,78,82</sup> | No consistent heterogeneity by HF history for SGLT2is; No analysis on heterogeneity by HF history performed for GLP1-RAs. |
| HF severity/score | 0 | 0 | 0 | 1<sup>100</sup> | 3<sup>157,180,181</sup> | 1<sup>74</sup> | Greater relative benefit of SGLT2i in those with NYHA class II vs class III/IV in one meta-analysis; No analysis of heterogeneity by HF severity/score performed for GLP1-RAs. |
| Renal function | 0 | 0 | 1<sup>91</sup> | 2<sup>100,101</sup> | 6<sup>13,114-116,171,177</sup> | 5<sup>2,74,78,79,82</sup> | No consistent heterogeneity by renal function. Single meta-analysis showed greater SGLT-2 benefit with lower eGFR and higher ACR; No evidence for heterogeneity by renal function for GLP1-RAs. |
| Genetics | 0 | 0 | 0 | 0 | 0 | 0 |  |
| Non-routine biomarkers | 0 | 0 | 0 | 0 | 6<sup>86,153,172-174,182</sup> | 0 | No heterogeneity across a variety of non-routine biomarkers. No analysis on heterogeneity by non-routine biomarkers performed for GLP1-RAs. |
| **Renal (eGFR changes / CKD progression / composite outcomes of these with or without** | **GRADE EVIDENCE B** |  |  | **GRADE EVIDENCE B** |  |  |  |
| **ACR changes)** |  |  |  |  |  |  |  |
| Baseline HBA1c | 0 | 0 | 0 | 0 | 0 | 0 |  |
| Renal Function | 0 | 3<sup>119,124,125</sup> | 1<sup>5</sup> | 2<sup>126,127</sup> | 6<sup>111,112,114-116,124</sup> | 5<sup>2,79,108-110</sup> | Generally no relationship between either eGFR or ACR and GLP1-RA benefit. Greater relative benefit of SGLT2i in those with higher eGFR (although inconsistent results with some studies showing no impact and 1 observational study finding the opposite relationship [Koh 2021]). Generally no relationship between ACR/proteinuria and SGLT2i benefit. |
| BMI | 0 | 2<sup>119,123</sup> | 0 | 1<sup>126</sup> | 2<sup>84,178</sup> | 1<sup>76</sup> | Greater GLP1-RA benefit with lower BMI but not seen consistently. Generally no effect on SGLT2i benefit |
| Age | 0 | 1<sup>118</sup> | 0 | 2<sup>126,127</sup> | 0 | 1<sup>76</sup> | No effect on GLP1-RA or SGLT2i benefit |
| Diabetes duration | 0 | 1<sup>121</sup> | 0 | 0 | 0 | 1<sup>76</sup> | No effect on GLP1-RA or SGLT2i benefit |
| Sex | 0 | 0 | 0 | 1<sup>126</sup> | 1<sup>167</sup> | 1<sup>76</sup> | No effect on SGLT2i benefit |
| Ethnicity | 0 | 0 | 0 | 0 | 2<sup>178,183</sup> | 1<sup>76</sup> | No effect on SGLT2i benefit |
| Genetics | 0 | 0 | 0 | 0 | 0 | 0 |  |
| Non-routine biomarkers | 0 | 0 | 0 | 0 | 5<sup>86,117,172-174</sup> | 0 | No effect on SGLT2i benefit |
| Blood pressure/hypertension | 0 | 2<sup>119,120</sup> | 0 | 1<sup>126</sup> | 0 | 1<sup>76</sup> | No effect on GLP1-RA or SGLT2i benefit |
| History of CVD/HF | 0 | 2<sup>119,122</sup> | 0 | 1<sup>126</sup> | 3<sup>162,163,184</sup> | 3<sup>2,77,109</sup> | No effect on GLP1-RA or SGLT2i benefit |
| **Renal (albuminuria changes)** | **GRADE EVIDENCE B** |  |  | **GRADE EVIDENCE B** |  |  |  |
| Baseline HBA1c | 0 | 0 | 0 | 0 | 0 | 0 |  |
| Renal Function | 0 | 2<sup>119,125</sup> | 1<sup>5</sup> | 0 | 0 | 1<sup>110</sup> | Greater GLP1-RA benefit with higher ACR although not seen consistently. No relationship between eGFR and GLP1-RA. No effect on SGLT2i benefit |
| BMI | 0 | 1<sup>119</sup> | 0 | 0 | 0 | 0 | No effect on GLP1-RA benefit |
| Age | 0 | 0 | 0 | 0 | 0 | 0 |  |
| Diabetes duration | 0 | 0 | 0 | 0 | 0 | 0 |  |
| Sex | 0 | 0 | 0 | 0 | 0 | 0 |  |
| Ethnicity | 0 | 0 | 0 | 0 | 1<sup>183</sup> | 0 | No effect on SGLT2i benefit |
| Genetics | 0 | 0 | 0 | 0 | 0 | 0 |  |
| Non-routine biomarkers | 0 | 0 | 0 | 0 | 1<sup>117</sup> | 0 | Single trial found greater SGLT2i benefit at higher IGFBP7 |
| Blood pressure/hypertension | 0 | 1<sup>119</sup> | 0 | 0 | 0 | 0 | No effect on GLP1-RA benefit |
| History of CVD/HF | 0 | 1<sup>119</sup> | 0 | 0 | 0 | 0 | No effect on GLP1-RA benefit |

A key finding across multiple studies including appropriately adjusted analysis of RCT and observational data was that HbA1c reduction with SGLT2i is substantially reduced with lower eGFR^13–19^. In pooled RCT data for canagliflozin 300mg, 6-month HbA1c reduction was estimated to be 11.0 mmol/mol for participants with eGFR ≥90 mL/min/1.73 m^2^, compared to 6.7 mmol/mol for those with eGFR 45-60^19^. With empagliflozin 25mg, 6-month HbA1c reduction was 9.6 mmol/mol at eGFR ≥90, and 4.3 mmol/mol at eGFR 30-60^16^.

A further finding confirmed by multiple robust studies is that in keeping with other glucose-lowering agents higher baseline HbA1c is associated with greater HbA1c lowering with SGLT2 inhibitors, including verses placebo^12,18, 20–24^. Active comparator studies suggested that higher baseline HbA1c may predict greater relative HbA1c response to SGLT2i therapy in comparison to DPP4i and sulfonylurea therapy^12,22,23^. Notably, an individual participant data meta-analysis of two RCTs showed greater improvement with empagliflozin (6-month HbA1c decline per unit higher baseline HbA1c [HbA1c slope] −0.49%] compared to sitagliptin (6-month HbA1c slope −0.29% [95%CI −0.42, −0.15]) and glimepiride (12-month HbA1c slope: empagliflozin −0.52% [95%CI −0.59, −0.44]; glimepiride −0.32% [95%CI −0.39, −0.25])^22^.

A number of studies assessing differences in glycaemic response to SGLT2i by ethnicity suggest that initial glycaemic response to this medication class does not vary by ethnicity^25–29^. Similarly, many studies also showed that response did not vary meaningfully vary by sex. Some studies suggested older age may be associated with reduced glycaemia response; however, analyses usually did not adjust for eGFR which may confound this association as eGFR declines with age^14,20, 29–32^.

#### GLP1-RA

Of 49 studies that met our inclusion/exclusion criteria, 24 observational studies, 6 post-hoc analysis of individual randomized control trials (RCTs), and 19 meta-analyses were included (**Supplementary Table 1f**). The majority of included studies assessed routine clinical characteristics and routinely measured clinical biomarkers, although 3 studies evaluated genetic variants, and 15 studies evaluated non-routine biomarkers (**Table 1**).

Studies consistently identified baseline HbA1c as a predictor of greater HbA1c response. For other clinical features, the strongest evidence was that in many observational studies markers of lower insulin secretion (including longer diabetes duration [or proxies such as insulin treatment], lower fasting C-peptide, lower urine C-peptide-to-creatinine ratio, and positive GAD or IA2 islet autoantibodies) were associated with lesser glycaemic response to GLP1-RA^33–46^. One large prospective study (n=620) observed clinically relevant reductions in HbA1c response with GLP1-RA in individuals with GAD or IA2 autoantibodies (mean HbA1c reduction −5.2 vs. −15.2 mmol/mol without autoantibodies) or C-peptide <0.25 nmol/L (mean HbA1c reduction −2.1 vs. −15.3 mmol/mol with C-peptide >0.25 nmol/L). In contrast, post-hoc RCT analyses has found T2D duration^47^ and beta-cell function^48,49^ do not modify glycaemic outcome. This may reflect trial inclusion criteria as included participants had relatively higher beta-cell function, and were less-commonly insulin-treated, compared with the observational cohorts^48^.

Few studies contrasted HbA1c outcome for GLP1-RA versus a comparator drug. One meta-analysis showed a greater HbA1c reduction with the GLP1-RA liraglutide compared to other antidiabetic drugs (sitagliptin, glimepiride, rosiglitazone, exenatide, and insulin glargine) across all baseline HbA1c categories (n=1,804)^50^, a finding supported for the GLP1-RA dulaglutide compared to glimepiride and insulin glargine^51^.

Overall, there was no consistent evidence for effect modification by body mass index (BMI), sex, age or kidney function, with studies reporting contrasting, or null, associations for these clinical features^36,37, 41–43,47, 51–61^. In comparative analysis, one large observational study found that markers of insulin resistance (including higher HOMA-IR, BMI, fasting triglycerides, and HDL) do not alter GLP1-RA response, but are associated with lesser DPP4-inhibitor response^54^.

There was limited evidence for differences by ethnicity. One large pooled RCT analysis (N=2,355) suggested greater HbA1c response in Asian participants compared to those of other ethnicities, but other studies have not identified differences in response across ethnic groups^62–65^. Similarly, limited studies evaluated pharmacogenetics, although two small studies suggest variants rs163184 and rs10305420, but not rs3765467, may be associated with lesser response in Chinese patients^40,66^.

### SGLT2i, GLP1-RA AND CARDIOVASCULAR OUTCOMES

#### SGLT2i: Evidence from clinical trials

Of 65 studies, 58 were post-hoc meta-analysis of RCTs or meta-analysis of multiple RCTs. Heart failure (HF) was common as a secondary outcome. The majority of studies were derived from EMPA-REG^67^ and the CANVAS program^68^, although more recent meta-analyses included up to 12 cardiovascular outcome trials (CVOTs) with different inclusion criteria, treatments, primary outcomes, and follow-up duration (**Supplementary Table 2a**). Most studies included only participants with established CVD or elevated cardiovascular risk, although some studies were restricted to patients with pre-existing heart failure or chronic kidney disease. While most CVOTs and meta-analyses included only patients with type 2 diabetes, some meta-analyses also included data from patients without diabetes in the EMPEROR-P^69^, EMPEROR-R^70^, DAPA-HF^71^ and DAPA-CKD^72^ RCTs. Studies primarily focused on relative rather than absolute treatment effects and one of two primary outcomes: 3-point MACE which was a composite of cardiovascular death, non-fatal MI, and non-fatal stroke; or composite heart failure outcomes including hospitalized heart failure and cardiovascular death. The longest duration of follow-up was in the CANVAS CVOT with a median follow-up of 5.7 years, although most other included CVOTs had durations of 1 to 4 years.

On average, in relative terms SGLT2i reduce the risk of cardiovascular disease (MACE) by 10% (HR 0.90 [95%CI 0.85, 0.95]), and heart failure hospitalization by 32% (HR 0.68 [95%CI 0.61, 0.76]) in individuals at with or at high-risk of CVD^2^. The majority of meta-analyses of CVOTs found no significant interactions for MACE or heart failure outcomes across a variety of biomarkers (**Supplementary Table 1a**). Several meta-analyses found no interactions by age, sex, and adiposity for MACE or heart failure outcomes. Four meta-analyses examined interactions by race for MACE outcomes and found no interactions. Three meta-analyses have consistently identified a greater relative heart failure benefit of SGLT2i in people of Black and Asian ethnicity^73–75^ (HR SGLT2i versus placebo 0.60 [95% CI 0.47, 0.74]) compared to White individuals (HR 0.82 [95% CI 0.73, 0.92])^73^, however one meta-analysis reported no difference between Caucasian and non-Caucasian individuals^76^.

Contemporary meta-analysis incorporating the CREDENCE and VERTIS-CV trials alongside EMPA-REG, CANVAS, and DECLARE suggests history of cardiovascular disease does not modify the efficacy of SGLT2i for MACE^2,77^. One meta-analysis suggests heart failure severity modifies the efficacy of SGLT2i’s for heart failure outcome (composite outcome of cardiovascular death or hospitalization for heart failure) with greater efficacy in patients with NYHA heart failure class II (HR SGLT2i versus placebo 0.66 [95%CI 0.59, 0.74]) than class III or IV (HR 0.86 [95%CI 0.75, 0.99])^74^. Other meta-analyses that examined treatment effect heterogeneity using heart failure history as a binary predictor did not find significant interactions^2,78^.

A recent meta-analysis^79^ that included 6 CVOTs of patients with diabetes and 4 CVOTs of patients with and without diabetes found that eGFR did not alter the relative benefit of SGLT2 inhibitors for MACE and heart failure outcomes^2,74,78, 80–82^; however, a greater relative benefit was reported for MACE in those with higher baseline albuminuria (ACR>300mg/g HR 0.74 95%CI 0.66, 0.84; ACR 30-300mg/g HR 0.95 [95%CI 0.82, 1.10]) ACR<30mg/g HR 0.87 [95%CI 0.77, 0.98]).

We identified many secondary analyses of single CVOTs, which largely found no interactions by biomarkers (**Supplementary Table 1a**). Single studies identified potential effect modification for MACE by history of CVD^83^, and obesity^84^, and history of heart failure for heart failure outcome^85^, but these associations were not replicated across the other studies or in multi-RCT meta-analysis. In a secondary analysis of CANVAS, participants with higher levels of biomarkers of cardiovascular stress (high-sensitivity cardiac troponin T (hs-cTnT), soluble suppression of tumorigenesis-2 (sST2), and insulin-like growth factor binding protein 7 (IGFBP7)) level had greater relative benefit for MACE; for a multimarker score summing high levels of these 3 biomarkers the relative benefit of SGLT2i for no abnormal biomarkers was HR: 0.99; 95% CI: 0.66-1.49, 1 abnormal biomarker (HR: 1.34; 95% CI: 0.94-1.89), 2 abnormal biomarkers (HR: 0.61; 95% CI: 0.45-0.82), and 3 abnormal biomarkers (HR: 0.46; 95% CI:0.18-1.17; P_interaction trend_ =0.005)^86^. Unlike meta-analyses, studies based on single RCTs typically performed multivariable adjustment for potential confounders.

#### *GLP1-RA:* Evidence from clinical trials

Of the 35 studies that investigated heterogeneity in effect of GLP1-RAs on cardiovascular health and met our inclusion criteria, 15 were meta-analyses of RCTs or pooled analyses of multiple RCTs, 15 were post-hoc analyses of RCTs, and 5 were observational studies (**Supplementary Table 1b**). Most studies used data collected from the LEADER, SUSTAIN 6, and EXSCEL trials, however in total the data from 7 CVOTs were used (**Supplementary Table 2b**). The majority of these CVOTs investigated the effect of previous cardiovascular disease on the cardiovascular efficacy of GLP1-RAs using 3-point MACE as a primary outcome, and with heart failure being a common secondary outcome, focusing on relative rather than absolute benefit. The population of 6 of the 7 CVOTs had established cardiovascular disease or high cardiovascular disease risk. The CVOT with the longest median follow up was REWIND with a median follow up of 5.4 years, and the median follow up of the other CVOTs ranged from 1 to 4 years.

Contemporary meta-analysis data suggests GLP1-RA reduce the relative risk of cardiovascular disease (MACE) by 14% (HR 0.86 [95%CI 0.80-0.93]), and heart failure hospitalization by 11% (HR 0.89 [95%CI 0.82, 0.98]) compared to placebo^3^. Several large meta-analyses examining heterogenous treatment effects in placebo-controlled CVOTs have been conducted for GLP1-RA^73,80,81, 87–94^, with the majority of studies focusing on whether prior established CVD modifies the relative effect of GLP1-RA on MACE and/or heart failure. Two meta-analyses reported the relative MACE benefit of GLP-RA may be restricted to those with established CVD^80,87^, the largest of which included 7 RCTs and reported a 14% relative risk reduction with GLP1-RA specific to individuals with established CVD (with CVD: HR 0.86 [95%CI 0.80, 0.93]; at high-risk of CVD: HR 0.94 [95% CI 0.82, 1.07])^80^. However, this risk difference is not conclusive and has not been replicated in other meta-analyses and pooled RCT analyses^88–90,95,96^, including an individual participant level re-analysis of the SUSTAIN and PIONEER RCTs which evaluated baseline CVD risk as a continuous rather than subgroup-level variable^97^.

Differential relative effects of GLP1-RAs on MACE have been reported by ethnicity in two out of three meta-analyses^73,80,87^: one showed a benefit of GLP1-RA treatment compared to placebo in Asian (HR 0.76 [95%CI 0.61, 0.96]) and Black (HR 0.77 [95%CI 0.59, 0.99]) individuals, but not in white individuals (HR 0.95 [95%CI 0.88, 1.02])^87^; the second another showed a significantly greater benefit of GLP1-RA for MACE in Asian compared to White individuals (HR Asian 0.68 [95%CI 0.53, 0.84]; White 0.87 [95% 0.81, 0.94])^73^. For other clinical features including sex, BMI/obesity, baseline kidney disease, duration of diabetes, baseline HbA1c, background glucose lowering medications, and prior history of microvascular disease, the overall body of evidence from meta-analyses does not provide robust evidence to support differential effects of GLP1-RA on CVD outcomes (**Table 1**).

#### SGLT2i and GLP1-RA: Evidence from observational studies

10 observational studies met our inclusion criteria, with studies primarily reporting relative rather than absolute risk differences^98–107^. These studies comparing SGLT2i and GLP1-RA individually with other oral therapies (predominantly DPP4i) generally reported average relative benefits for CVD and heart failure outcomes in-line with placebo-controlled trials, with no consistent pattern of subgroup level differences across studies (**Supplementary Table 1a and 1b**).

A few observational studies compared SGLT2i and GLP1-RA CVD outcomes. In a US claims-based study with follow-up to two years (n=47,343), Htoo et al.^103^ reported a higher relative risk of MACE with SGLT2i compared to GLP1-RA specific to individuals without CVD and heart failure (Relative risk [RR] 1.31 [95% CI 1.09, 1.56]), and a higher risk of stroke with SGLT2i versus GLP1-RA specific to individuals without CVD (No CVD without heart failure: RR 1.62 [95%CI 1.10, 2.38]; No CVD with heart failure: RR 3.30 [95%CI 1.22, 8.97]). In contrast, over a median follow-up of 7 months Patorno et al.^102^ reported a lower relative risk of myocardial infarction with SGLT2i compared to GLP1-RA in US claims data specific to individuals with a history of CVD (n=156,825; HR 0.82 [95%CI 0.71, 0.95]), with no differences in stroke outcomes irrespective of CVD status. Both studies reported a consistent benefit of SGLT2i over GLP1-RA for heart failure. Raparelli et al.^99^ identified potential differences by sex in the Truven Health MarketScan database (n=167,341), with, compared to sulfonylureas and over a median follow-up of 4.5 years, a greater relative reduction with GLP1-RA for females (HR 0.57 [95%CI 0.48, 0.68]) compared to males (HR 0.82 [95%CI 0.71, 0.95]), but a similar benefit for both sexes with SGLT2i (females HR 0.58 [95%CI 0.57, 0.83]; males HR 0.69 [95%CI 0.57, 0.83]).

### SGLT2i, GLP1-RA AND RENAL OUTCOMES

#### SGLT2i: Evidence from clinical trials

29 studies met our inclusion criteria. These included 2 analyses of observational data, 20 post-hoc analyses of individual RCTs, and 7 trial meta-analyses (**Supplementary Table 1c**). All of the post-hoc RCT analyses and all but 1 of the meta-analyses used only data from the 12 SGLT2i cardiovascular/renal RCTs shown in Supplementary Table 2a, which therefore provided most of the evidence in our review. These trials included people with type 2 diabetes with and without pre-existing cardiovascular disease, and had composite renal endpoints incorporating two or more of the following: changes in eGFR/serum creatinine, end-stage renal disease, changes in urine albumin:creatinine ratio (ACR), and/or death from renal causes; which differed between trials. Most studies assessed routine clinical characteristics, especially renal function as measured by eGFR or urine ACR or a combination of both. In addition, 4 post-hoc RCT analyses examined non-routine plasma biomarkers. We found no genetic studies.

On average, SGLT2i have a relative benefit for a number of renal outcomes including kidney disease progression (HR 0.63, 95%CI 0.58,0.69) and acute kidney injury (HR 0.77, 95%CI 0.70, 0.84)^4^. Placebo-controlled trial meta-analyses of subgroups found no evidence for heterogeneity of SGLT2i treatment effects on relative renal outcomes by age^76^, use of other glucose-lowering drugs^76^, use of blood pressure/cardiovascular medications^76,108^, blood pressure^76^, BMI^76^, diabetes duration^76^, Caucasian ethnicity^76^, history of cardiovascular disease or heart failure^2,77^ or sex^76^.

For baseline eGFR, an early meta-analysis that included EMPA-REG, CANVAS and DECLARE reported greater effect of SGLT2i on renal outcomes in those with higher eGFR^109^ but both a later meta-analysis that added CREDENCE^108^ and a recent meta-analysis that added two further studies (SCORED and DAPA-CKD, including some participants without diabetes)^79^ showed no effect of baseline eGFR on renal outcomes with SGLT2i. For urine ACR, meta-analyses of subgroups found no evidence for greater SGLT2i effect with higher UACR^2,79,108,110^. Single RCTs found no heterogeneity of treatment effect by eGFR and UACR, or subgroups defined by the combination of these two^111–115^, with the except of Neuen et al.^116^ which showed a greater SGLT2i effect in preventing eGFR decline relative to placebo for those with higher UACR, and heterogeneity in a composite renal outcome by UACR. Overall there was limited or no evidence to support modifying effects of baseline eGFR or UACR on the effect of SGTL2i on renal function outcomes.

A few post-hoc analyses of the CANVAS RCT considered non-routine biomarkers, with most showing no interaction with SGLT2i treatment and renal outcomes. Two RCTs studied the effect of SGLT2i on renal outcomes at differing plasma IGFBP7 levels. One study reported an interaction of IGFBP7 with SGLT2i treatment for progression of albuminuria (>96.5ng/ml HR 0.64; <=96.5ng/ml HR 0.95, P*_interaction_* = 0.003)^117^ but no effect was seen for the composite renal endpoint in two studies^86,117^. The biomarker panel (sST2, IGFBP7, hs-cTnT) that showed a strong interaction with SGLT2i for MACE outcomes (see above) did not show any interaction for renal outcomes^86^.

#### GLP1-RA: Evidence from clinical trials

7 studies met our inclusion criteria: all post-hoc RCT analyses, 6 of individual trials (or multiple trials analysed separately) and 1 pooled analysis of two RCTS (**Supplementary Table 1d**). These studies used data from 5 of the 7 GLP1-RA cardiovascular outcome trials shown in Supplementary Table 2b, with renal outcomes only a secondary endpoint. Most of these trials had composite renal endpoints as per the SGLT2i cardiovascular/renal trials, whilst some examined changes in either eGFR or urine ACR only. All studies assessed routine clinical characteristics, especially renal function as measured by eGFR or urine ACR. No studies of genetics or non-routine biomarkers were identified. The overall sample sizes were small and subgroup analyses underpowered to show a subgroup by treatment interaction for renal outcomes.

Overall, GLP1-RA reduce the relative risk of albuminuria over 2 years by 24% versus placebo (HR 0.76 [95% CI 0.73-0.80; P<0.001), and similarly reduce the relative risk of a 40% reduction in eGFR (HR, 0.86 [95% CI 0.75-0.99]; P=0.039)^5^. Studies found no heterogeneity of GLP1-RA relative treatment effect by age^118^, blood pressure^119,120^, diabetes duration^121^, history of cardiovascular disease/heart failure^119,122^ or use of RAS inhibitors^119^. For BMI, a post-hoc analysis of EXSCEL (Exenatide) found a greater GLP1-RA effect on reducing rate of eGFR decline in those with lower BMI (BMI≤30kg/m^2^ treatment difference 0.26 mL/min/1.73m^2^/year [95% CI 0.04, 0.48] vs BMI>30 kg/m^2^ −0.12 [−0.26, 0.03], P_interaction_ =0.005)^119^. However, Verma et al.^123^ found no significant interaction by BMI subgroup with GLP1-RA treatment for a composite renal outcome in LEADER (Liraglutide) or SUSTAIN 6 (Semaglutide).

For baseline eGFR, a pooled analysis of LEADER and SUSTAIN-6 reported a significant interaction, with lower eGFR associated with greater GLP1-RA effect in reducing eGFR decline: Semaglutide 1.0mg vs placebo, eGFR<60 difference in decline 1.62 ml/min/1.73m^2^/year vs eGFR>=60 difference in decline 0.64ml/min/1.73m^2^/year, P_interaction_=0.057; Liraglutide 1.8mg vs placebo, eGFR<60 difference in decline 0.67 ml/min/1.73m^2^/year vs 0.15ml/min/1.73m^2^/year, P_interaction_=0.008)^5^. However, a study of Exenatide LAR found no treatment heterogeneity for this same outcome by eGFR category^119^ and in a further analysis of LEADER the renal composite endpoint was used with no interaction reported by baseline eGFR category^124^. The overall evidence does not support an effect of baseline eGFR on the relative renal benefit for GLP1-RA as an overall drug class.

For baseline UACR, a pooled analysis of LEADER and SUSTAIN-6^5^ and EXSCEL^119^ showed a greater benefit of GLP1-RA on eGFR reduction or eGFR slope with higher UACR however there was either no significant interaction^5^ or no formal interaction test was reported^119^. For ELIXA, Muskiet et al.^125^ did not find a significant interaction of UACR category on eGFR decline. A further study found no association between UACR and GLP1-RA effect on reducing a composite renal outcome^124^.

Two studies found that GLP1-RAs more effectively reduced UACR in those with higher UACR. In a pooled analysis of LEADER and SUSTAIN-6, those with normalbuminuria had a 20% (95%CI 15%, 25%) reduction in UACR compared to placebo; those with microalbuminuria had a 31% (95%CI 25-37%) reduction; those with macroalbuminuria had a 19% (95%CI 7-30%) P_interaction_=0.021^5^. In ELIXA, least-squares mean percentage change in UACR was –1·69% (SE 5·10; 95% CI –11·69 to 8·30; p=0·7398) in participants with normoalbuminuria, –21·10% (10·79; –42·25 to 0·04; p=0·0502) in participants with microalbuminuria, and –39·18% (14·97; –68·53 to –9·84; p=0·0070) in participants with macroalbuminuria in favour of lixisenatide; a formal test for interaction was not reported^125^. A third study found no treatment heterogeneity for this same outcome^119^.

In summary, the included studies showed conflicting results for renal outcomes of GLP1-RA, though the majority were underpowered to detect heterogenous treatment effects. The most consistent finding was that a higher UACR is associated with greater GLP1-RA reduction in UACR relative to placebo, but this does not translate to benefits in eGFR defined measures of renal function. There were no other biomarkers that robustly predicted benefit from GLP1-RA for the renal outcomes examined.

#### SGLT2i and GLP1-RA: Evidence from observational studies

There were no observational studies for GLP1-RA and renal outcomes included, and no comparison studies between people treated with GLP1-RA and SGLT2i. Observational studies comparing SGLT2i to other glucose-lowering drugs confirmed the lack of treatment effect heterogeneity associated with age^126,127^, use of blood pressure/cardiovascular medications^127^, blood pressure (Koh 2021), history of cardiovascular disease^126^ and sex^126^, but one study in a Korean population found greater SGLT2i benefit on progression to end stage renal impairment with higher BMI (BMI<25kg/m2, HR 0.80 (95%CI 0.51, 1.25); BMI≥25kg/m2 HR 0.27 (0.16, 0.44), P_interaction_=0.002) and with abdominal obesity compared to without^126^. This is not consistent with results from meta-analysis of RCTs.

### SUMMARY OF QUALITY ASSESSMENT

To evaluate risk of bias, we used the JBI critical appraisal tool for cohort studies as the best flexible tool for the range of studies included. Due to our screening criteria no manuscripts that passed full text screening were excluded due to risk of bias. The checklist results for the 11 points in the appraisal checklist are shown as a heatmap in **Supplementary Figure 2a (SGLT2i)** and **2b (GLP1-RA)**.

Additionally, the Grading of Recommendations, Assessment, Development, and Evaluations (GRADE) framework was applied at the outcome level for each drug class to determine the quality of evidence and certainty of effects for these subsections. For SGLT2i Cardiovascular (GRADE B), SGLT2i renal (GRADE B), GLP1-RA Cardiovascular (GRADE B) and GLP1-RA Renal (GRADE B), the majority of our evidence base comes from industry-funded, CVOTs (RCT designs), including post-hoc analyses of individual trials as well as meta-analyses. There were limited observational studies.

For the two glycaemia outcomes, we scored the SGLT2i glycaemia outcomes as GRADE B but the GLP1-RA as GRADE C. This reflects that a larger proportion of the studies included for evaluation of GLP1-RA outcomes were observational (24/49). By contrast, for SGLT2i glycaemia outcomes there were (18) RCT/meta-analyses and (9) observational studies.

## DISCUSSION

This systematic review provides a comprehensive review of observational and RCT-based studies of people with type 2 diabetes specifically examining heterologous treatment effects for SGLT2i and GLP1-RA therapies on glycaemic, cardiovascular, and renal outcomes. We assessed evidence for treatment effect modification for a wide range of demographic, clinical and biological features, including pharmacogenetic markers. Each of the three clinical outcomes were evaluated separately for each drug class for a total of 6 sub-studies. Overall, our review identified limited evidence for treatment effect heterogeneity for glycaemia, cardiovascular, and renal outcomes for the two drug classes. We summarize the key findings below.

For glycaemic response, there was strong evidence that reduced renal function is associated with lower efficacy of SGLT2i. For GLP1-RA, markers of reduced insulin secretion, either directly measured (e.g. c-peptide or HOMA-B) or proxy measures such as diabetes duration, were associated with reduced glycaemic response to GLP1-RA in the majority of observational studies. As with other glucose-lowering drug classes, a greater glycaemic response with both SGLT2i and GLP1-RA was seen at higher baseline HbA1c. We did not identify any studies examining whether the relative efficacy of SGLT2i compared to GLP1-RA is altered by baseline HbA1c levels.

For both CVD and heart failure outcomes, RCT meta-analyses do not support differences in the relative efficacy of either GLP1-RA or SGLT2i based on an individuals’ prior CVD status. However, this finding should be interpreted cautiously as all RCTs to-date have predominantly included participants with, or at high-risk of, CVD, thereby excluding the majority of the wider T2D population at lower risk. However, meta-analyses suggest the relative effects of both drug classes may be greater in people of non-white ethnicity. In particular, those of Asian and African ethnicity (compared to White Europeans) have been shown to have a greater relative benefit for hospitalization for heart failure/CV death (but not MACE) with SGLT2i, and MACE for GLP1-RA.

When evaluating renal outcomes, there was no consistent evidence of treatment heterogeneity for SGLT2i, but for GLP1-RA there was greater reduction in proteinuria in those with higher baseline proteinuria.

This limited evidence could reflect a true lack of heterogenous treatment effects, but is more likely to reflect an absence of clinical studies that were well designed or sufficiently powered to robustly identify and characterise treatment effect heterogeneity. Although five of the six sub-studies we evaluated were evaluated at GRADE B, there were methodological concerns with many of the included studies. As individual RCTs are by design powered only for the main effect of treatment^128^, our primary focus when reporting were meta-analyses of post-hoc subgroup analyses of RCTs, however we found the subgroup analyses in these studies primarily focused on stratification by baseline risk for the outcome in question e.g. baseline HbA1c on glycaemic response, CKD stage or albuminuria on renal outcomes, and CVD risk or established CVD for CVD outcomes. Other common subgroups included those defined by BMI, age, sex or other routinely collected clinical characteristics, with very few studies evaluating non-routine biomarkers or pharmacogenetic markers (as highlighted in **Table 1**). A major limitation was that studies predominantly focused on conventional “one-at-a-time” approaches to subgroup analysis, with very few studies assessing continuous features (such as BMI) on a continuous scale which is required to maximize power to detect treatment effect heterogeneity^128,129^.

It is also important to recognize that almost all the studies evaluating cardiovascular and renal endpoints included in our systematic review focused on the *relative* effect of a biomarker/stratifier on the outcome, as most studies reported a hazard ratio compared with a placebo arm for the outcome of interest (e.g. MACE, incident renal disease). This does not recognize that baseline absolute risk of these endpoints is likely to differ significantly across these strata; so although, for example, there was no difference in relative benefit of an SGLT2i by age, this means that on the absolute scale benefit will increase with age (as underlying absolute risk increases) and it is this absolute benefit that should be considered when deciding on whether to initiate SGLT2i treatment.

A significant finding of our review is the lack of robust comparative effectiveness studies directly examining treatment effect heterogeneity for these two major drug classes, either head-to-head or compared with other major anti-hyperglycaemic therapies. Insight into effect modification for a single drug class is not sufficient to support clinical translation of a precision medicine approach. The lack of direct comparisons between therapies obscures the interpretation of biomarkers with regards to whether they function as broad prognostic factors, which may be relevant to any (or at least multiple) drug class, or as markers of heterogenous treatment effects specific to a particular drug class. An evidence base that includes more high-quality studies on heterogeneity in the comparative effectiveness of SGLT2i, GLP1-RA, and other drug classes is needed to advance the field towards clinically useful precision diabetes medicine. For cardiovascular and renal outcomes, these studies need to incorporate both absolute outcome risk and relative estimates of treatment effects in order to usefully inform clinical decision making. Only when this evidence is available can precision medicine support more individualised treatment decisions, allowing providers to select an optimal therapy from a set of multiple options informed by each medication’s risk/benefit profile specific to the characteristics of an individual patient.

*We identified the following additional, high-level gaps in our review:*

- Limited head-to-head comparative effectiveness studies examining treatment effect heterogeneity.
- A lack of robust studies integrating multiple clinical features and biomarkers. The majority of studies only tested single biomarkers in “one-at-a-time” subgroup analysis.
- Few studies focused on pharmacogenetics or non-routine biomarkers.
- Few studies conducted in low-middle income countries, required for an equitable global approach to precision type 2 diabetes medicine.
- Few RCT meta-analyses based on individual-level participant data, precluding robust evaluation of between-trial heterogeneity and individual-level confounders.
- An absence of confirmatory studies. We identified no prospective studies testing *a priori* hypotheses of potential treatment effect modifiers, or studies conducting independent validation of previously described heterogenous treatment effects.
- A lack of population-based data representing individuals treated in routine care. As cardiovascular and renal trials have focused on high-risk participants, the benefits of SGLT2i and GLP1-RA for primary prevention is a major unanswered question.
- Few cardiovascular and renal outcome studies considering treatment effect modification on the absolute as well as relative risk scale.
- A focus on short-term glycaemic outcomes, with limited studies investigating durability of glycaemic response or time to glycaemic failure.

Of note, several studies published since our data extraction was completed in February 2022 are worth highlighting as they fill some of these evidence gaps: the TriMaster study – an ‘n-of-1’ precision medicine RCT of SGLT2i, DPP4i and TZD that established that individuals with higher renal function (eGFR >90 ml/min/1.73 m^2^) have a greater HbA1c response with SGLT2i vs DPP4i relative to those with eGFR 60–90 ml/min/1.73 m^2^ ^130^; a similarly designed two-way crossover trial in New Zealand which identified a greater relative benefit of TZD therapy compared to DPP4i in people with obesity and/or hypertriglyceridemia^131^; a study using large-scale observational data and post-hoc analysis of individual participant-level data from 14 RCTs that specifically investigated differential treatment effects with SGLT2i and DPP4i, and developed a treatment selection model to predict HbA1c response on the two therapies based on an individuals’ routine clinical characteristics^132^; and a robust study across observational and multiple RCTs identifying pharmacogenetic markers of differential glycaemic response to GLP1-RA^133^.

This review highlights the need for several research priorities to advance our limited understanding of heterogenous treatment effects among individuals with type 2 diabetes. We outline priorities for research to advance the field towards a translational model of evidence-based, empirical precision diabetes medicine (**Figure 1**), and highlight the recent Predictive Approaches to Treatment effect Heterogeneity (PATH) Statement to guide this research^129^. In the future, with a greater understanding of heterogenous treatment effects and enhanced capacity to predict individual treatment responses, precision treatment in type 2 diabetes may be able to integrate demographic, clinical, biological or other patient-level features to match individuals to their optimal anti-hyperglycaemic regimen.

**Figure 1.**
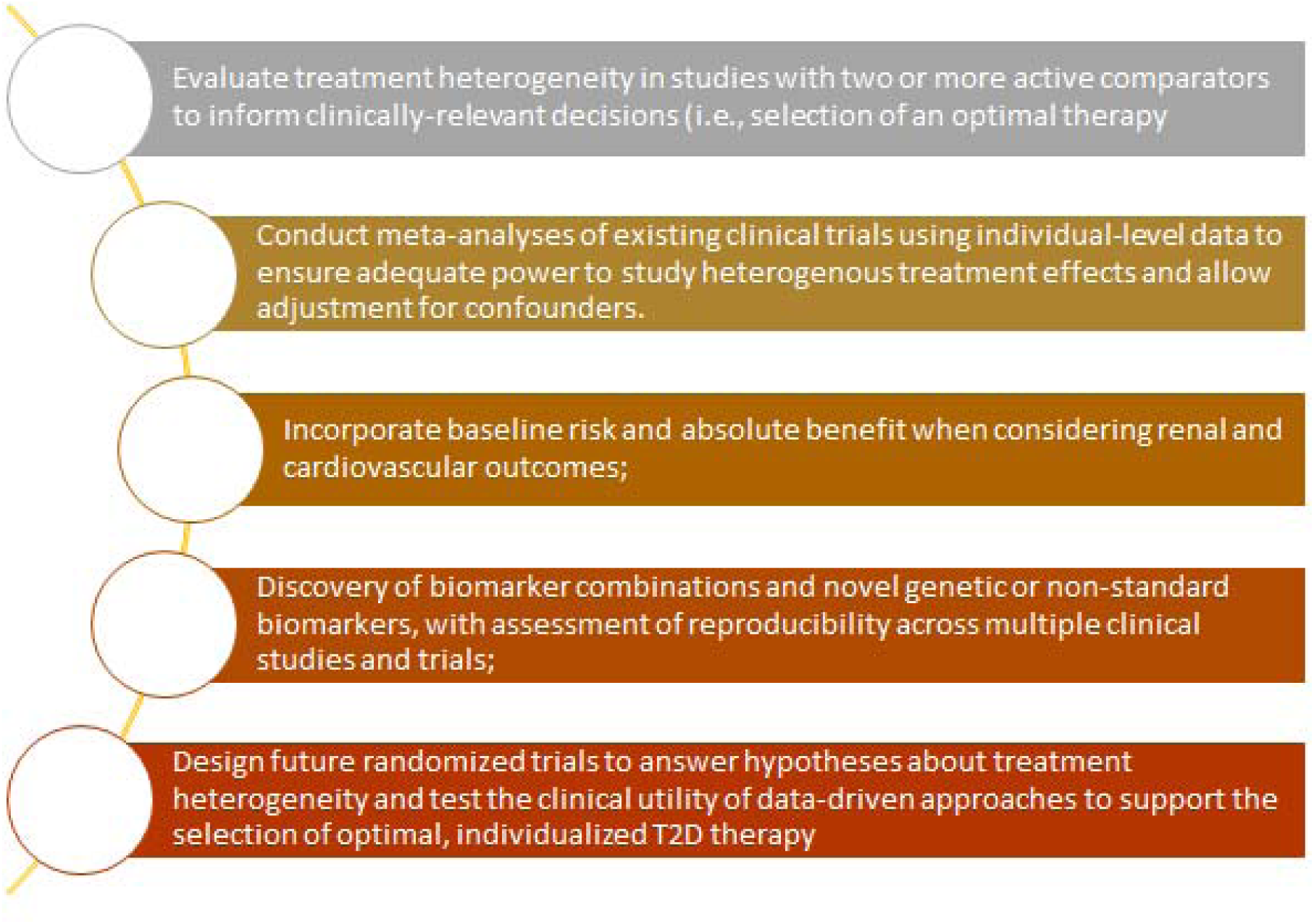
Priorities for future research to advance the field towards a translational model of evidence-based, empirical precision diabetes medicine

## Conclusion

There is limited evidence of treatment effect heterogeneity with SGLT2i and GLP1-RA for glycaemic, cardiovascular and renal outcomes in people with type 2 diabetes. This lack of evidence likely reflects the methodological limitations of the current evidence base. Robust future studies to fill the research gaps identified in this review are required for precision medicine in type 2 diabetes to translate to clinical care.

## Data Availability

Template data collection forms and the data extracted from included studies are available upon request.

## Acknowledgements

The ADA/EASD Precision Diabetes Medicine Initiative, within which this work was conducted, has received the following support: The Covidance license was funded by Lund University (Sweden) for which technical support was provided by Maria Björklund and Krister Aronsson (Faculty of Medicine Library, Lund University, Sweden). Administrative support was provided by Lund University (Malmö, Sweden), University of Chicago (IL, USA), and the American Diabetes Association (Washington D.C., USA). The Novo Nordisk Foundation (Hellerup, Denmark) provided grant support for in-person writing group meetings (PI: L Phillipson, University of Chicago, IL). This study was supported by the National Institute for Health and Care Research Exeter Biomedical Research Centre. The views expressed are those of the author(s) and not necessarily those of the NIHR or the Department of Health and Social Care.

## Funding

KGY and JMD are supported by Research England’s Expanding Excellence in England (E3) fund. ARK is supported by the National Center for Advancing Translational Sciences, National Institutes of Health, through Grant KL2TR002490. The content is solely the responsibility of the authors and does not necessarily represent the official views of the NIH. SR is funded by an US Department of Veterans Affairs Award IK2-CX001907, and a Webb-Waring Biomedical Research Award from the Boettcher Foundation. JMD is funded by the EFSD Rising Star Fellowship Programme, the UK Medical Research Council (MR/N00633X/1), and a BHF-Turing Cardiovascular Data Science Award (SP/19/6/34809). The funders had no role in the study design, data extraction or interpretation, writing of the article, or the decision to submit for publication.

## Conflict of interest

APM declares previous research funding from Eli Lilly and Company, Pfizer, and AstraZeneca. ERP has received honoraria for speaking from Lilly, Novo Nordisk and Illumina. All other authors declare no conflict of interest.

## Appendix 1: PRISMA CHECKLIST

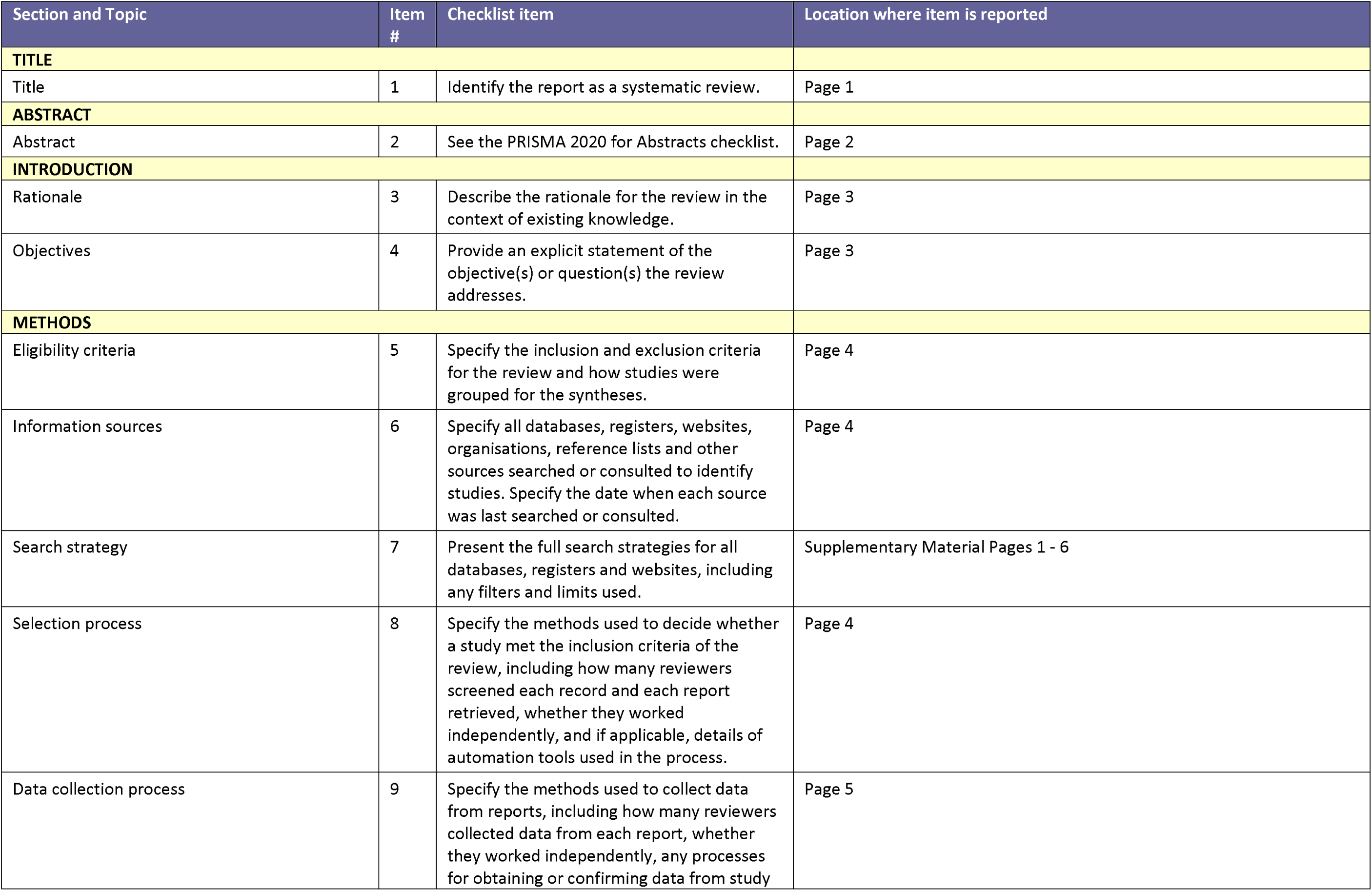
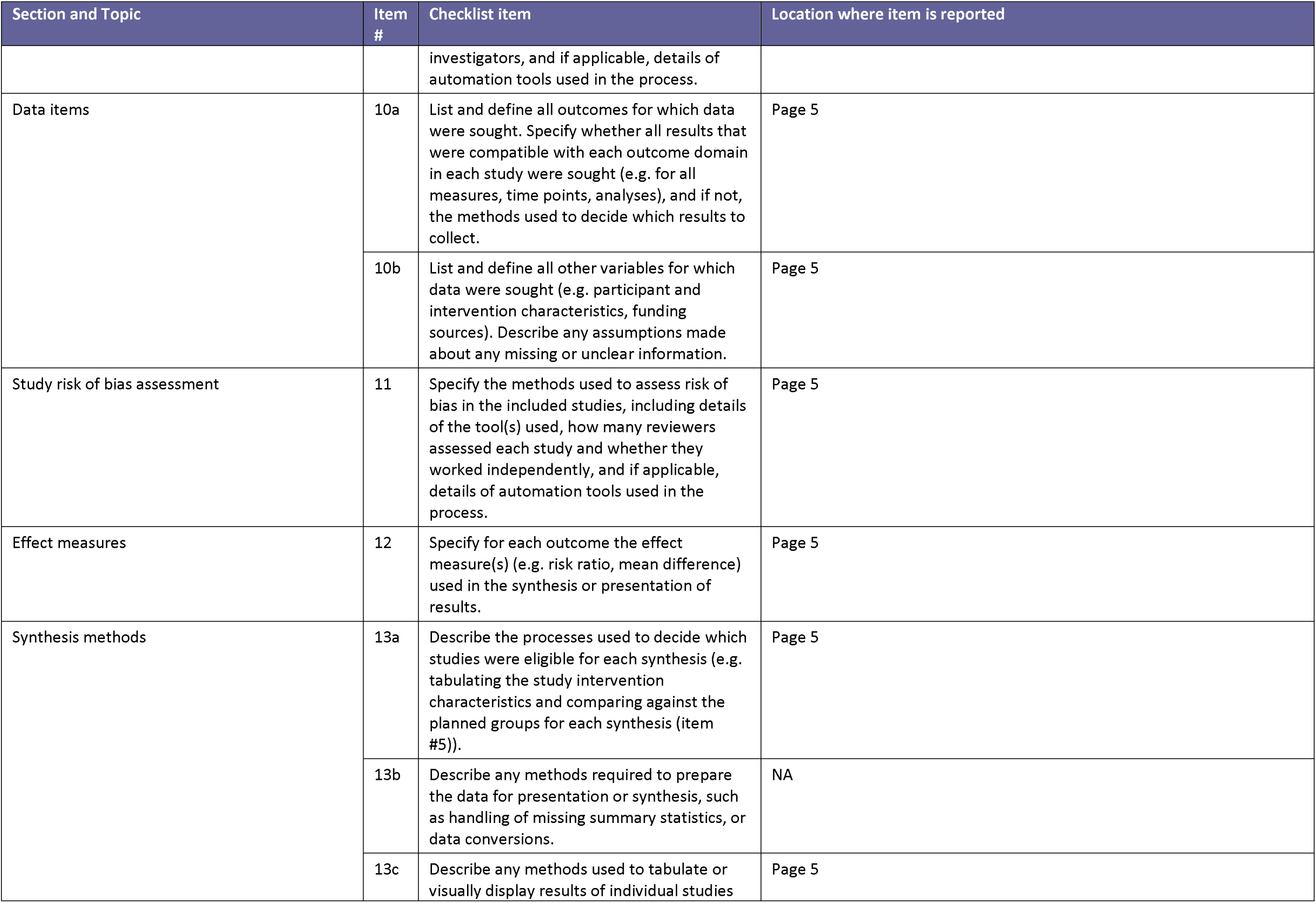
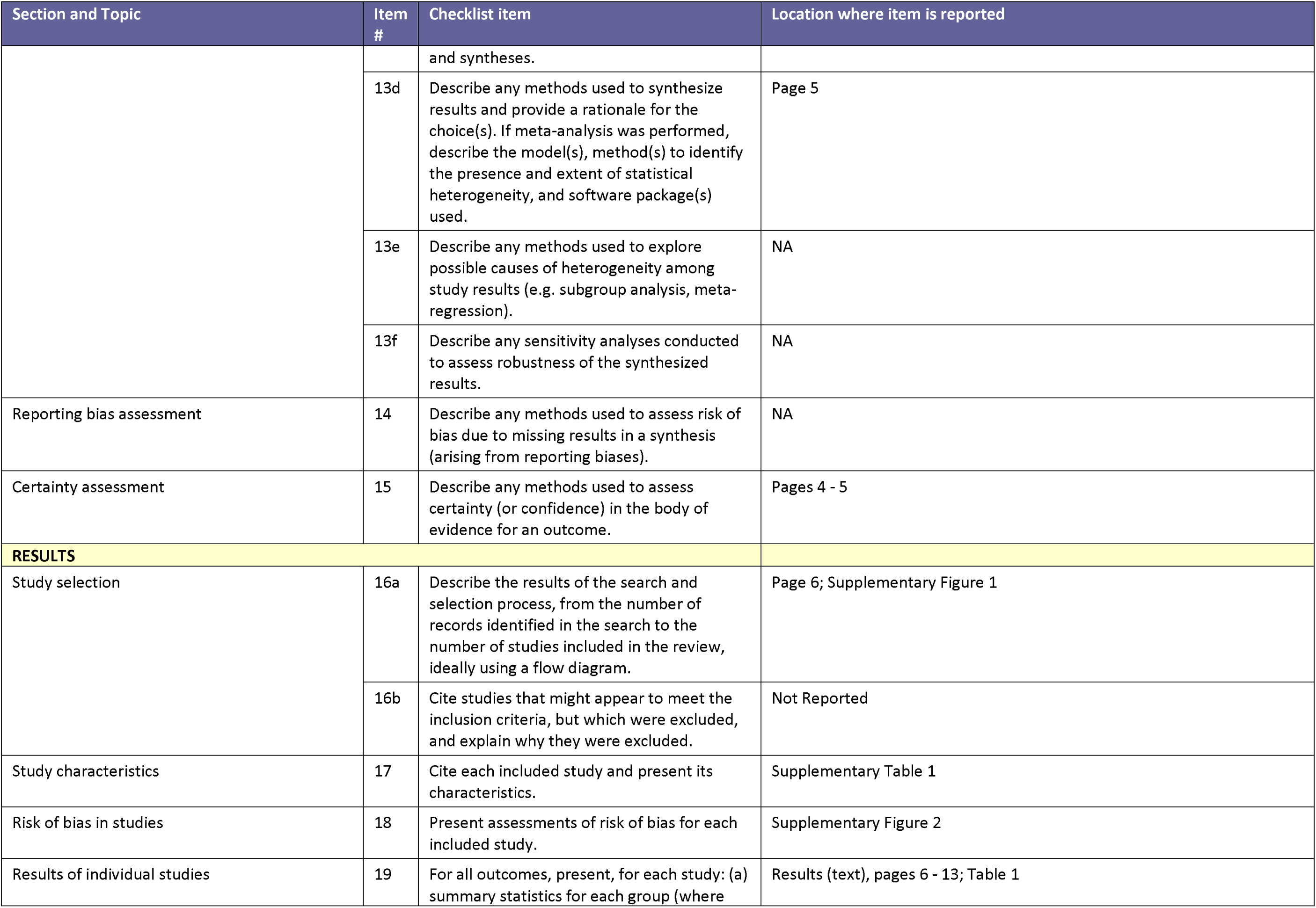
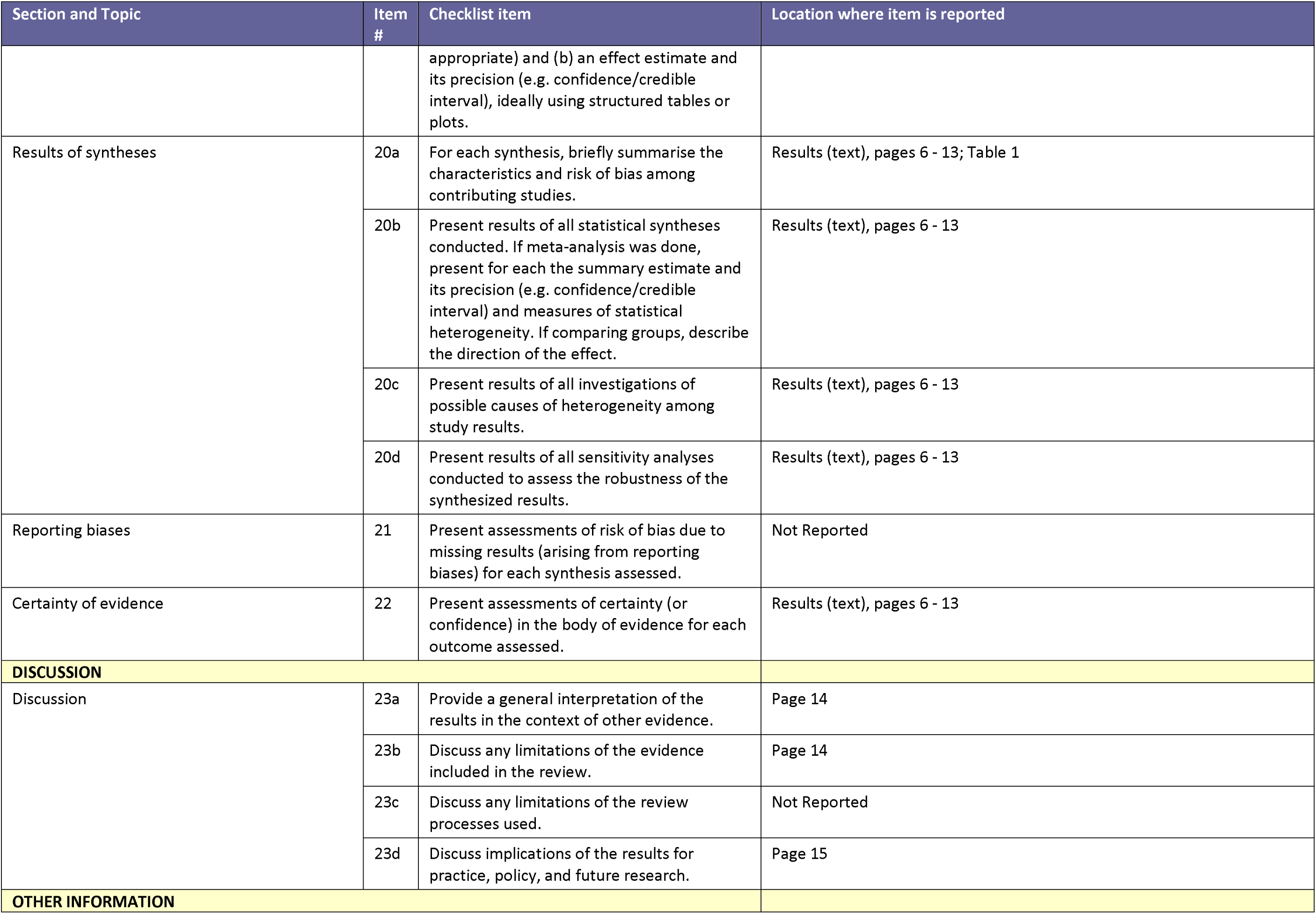
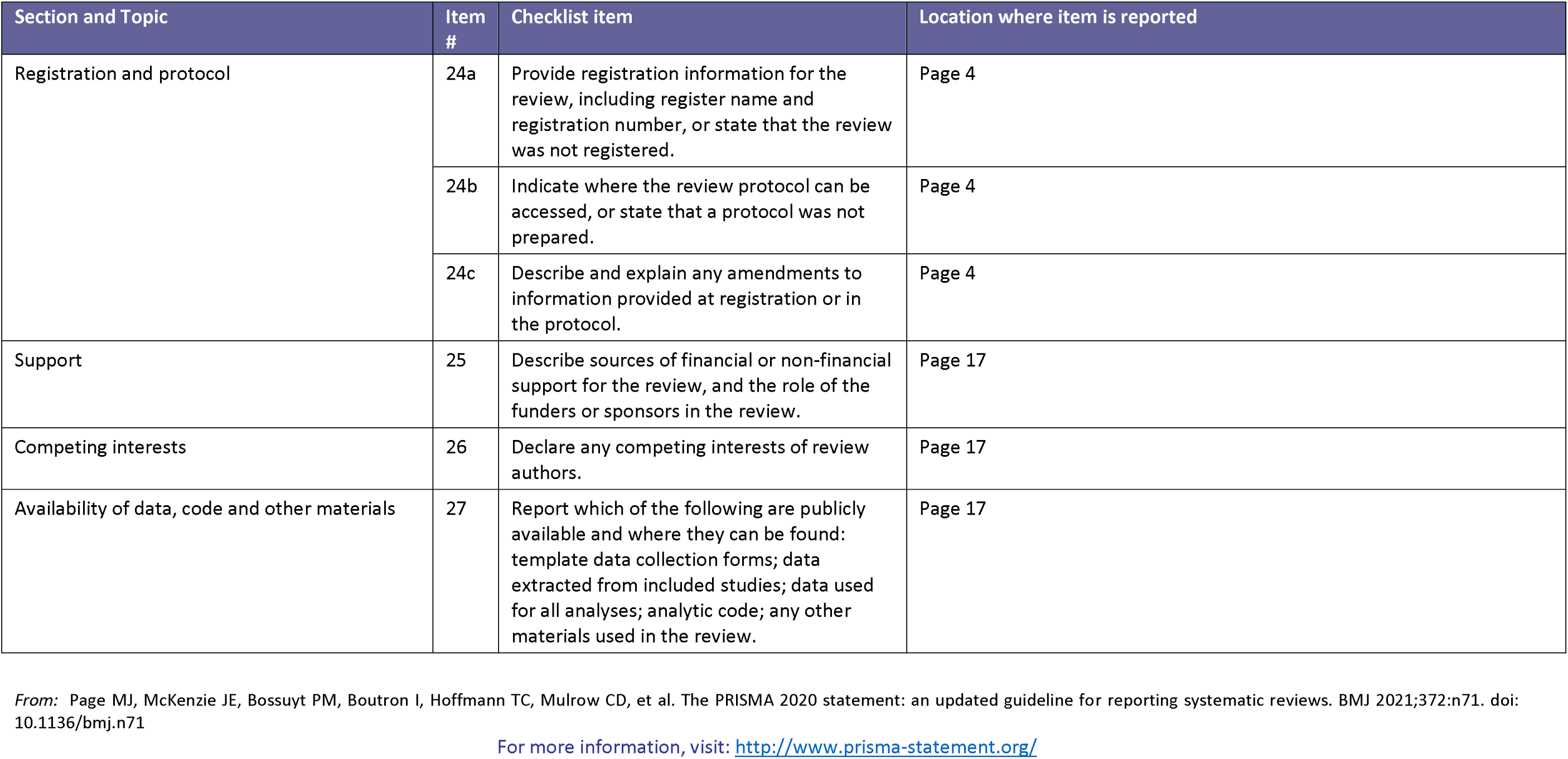

## Notes

### Clinical Protocols

https://www.crd.york.ac.uk/prospero/display_record.php?RecordID=303236

### Funding Statement

KGY and JMD are supported by Research Englands Expanding Excellence in England (E3) fund. ARK is supported by the National Center for Advancing Translational Sciences, National Institutes of Health, through Grant KL2TR002490. The content is solely the responsibility of the authors and does not necessarily represent the official views of the NIH. SR is funded by an US Department of Veterans Affairs Award IK2-CX001907, and a Webb-Waring Biomedical Research Award from the Boettcher Foundation. JMD is funded by the EFSD Rising Star Fellowship Programme, the UK Medical Research Council (MR/N00633X/1), and a BHF-Turing Cardiovascular Data Science Award (SP/19/6/34809). The funders had no role in the study design, data extraction or interpretation, writing of the article, or the decision to submit for publication.

