## Supplementary Material for "Precision medicine in type 2 diabetes: A systematic review of treatment effect heterogeneity for GLP1-receptor agonists and SGLT2-inhibitors"

#### 1. Search terms

##### GLP1RA

#1

"Diabetes Mellitus, Type 2"[Mesh] OR TD2[Title/Abstract] OR "Type 2 diabetes"[Title/Abstract] OR "Diabetes, type 2"[Title/Abstract

#2

GLP1RA OR "GLP1 Receptor Agonists" OR "GLP-1 Receptor Agonists" OR "GLP1RA" OR Exenatide OR Liraglutide OR Lixisenatide OR Semaglutide OR Dulaglutide OR Albiglutide

#3

White[Title/Abstract] OR Caucasian[Title/Abstract] OR Asian[Title/Abstract] OR African[Title/Abstract] OR Ethnicity[Title/Abstract] OR Ethnic[Title/Abstract]

#4

Age[Title/Abstract] OR "Diabetes duration"[Title/Abstract] OR "BMI"[Title/Abstract] OR "Body Mass Index"[Title/Abstract] OR Adiposity[Title/Abstract] OR "Sex"[Title/Abstract] OR "clinical features"[Title/Abstract]

#5

"Insulin resistant"[Title/Abstract] OR "Insulin deficient"[Title/Abstract] OR "beta-cell function"[Title/Abstract] OR "c-peptide*"[Title/Abstract] OR "metabolite*"[Title/Abstract] OR "Metabolomic"[Title/Abstract] OR "Proteomic"[Title/Abstract] OR "Protein*"[Title/Abstract] OR "peptide*"[Title/Abstract]

#6

(Genotype[Title/Abstract] OR SNP[Title/Abstract] OR variant[Title/Abstract] OR GWAS[Title/Abstract] OR "Genome Wide Association Study"[Title/Abstract] OR Genetic[Title/Abstract] OR Epigenetic[Title/Abstract] OR Methylation[Title/Abstract]) OR ((("Genotype"[Mesh]) OR "Genome-Wide Association Study"[Mesh]) OR "Methylation"[Mesh])

#7

Precision[Title/Abstract] OR Prediction[Title/Abstract] OR Stratification[Title/Abstract] OR Stratified [Title/Abstract] OR Individualised[Title/Abstract] OR Individualisation[Title/Abstract] OR Personalized[Title/Abstract] OR Personalization[Title/Abstract] OR targeted[Title/Abstract]

#8

RCT OR randomis* OR randomiz*

#9

#3 OR #4 OR #5 OR #6 OR #7

**OUTCOME: HBA1C**

#10

"hba1c reduction"[Title/Abstract] OR "hba1c change*"[Title/Abstract] OR "hba1c response"[Title/Abstract] OR "treatment response"[Title/Abstract] OR "glycaemic efficacy"[Title/Abstract] OR "glycemic efficacy"[Title/Abstract] OR fall in hba1c[Title/Abstract] OR change from baseline in hba1c[Title/Abstract] OR change in hba1c[Title/Abstract] OR changes in hba1c[Title/Abstract] OR slope of change[Title/Abstract] OR "hba1c outcome*"[Title/Abstract] OR outcome hba1c[Title/Abstract] OR "A1C change*"[Title/Abstract] OR "A1C reduction"[Title/Abstract] OR A1C response[Title/Abstract] OR A1C outcome*[Title/Abstract]

#11

#1 AND #2 AND #9 AND #10

**OUTCOME: CARDIOVASCULAR DISEASE**

#12

((((((((("Cardiovascular Diseases"[Mesh]) OR "Myocardial Infarction"[Mesh]) OR "Myocardial Revascularization"[Mesh]) OR "Acute Coronary Syndrome"[Mesh]) OR "Myocardial Ischemia"[Mesh]) OR "Heart Failure"[Mesh]) OR "Angioplasty"[Mesh]) OR "Percutaneous Coronary Intervention"[Mesh]) OR "Stroke"[Mesh]) OR "Angina Pectoris"[Mesh])

#13

"cardiovascular disease"[Title/Abstract] OR "myocardial infarction"[Title/Abstract] OR revasculari*[Title/Abstract] OR "acute coronary syndrome"[Title/Abstract] OR "coronary heart disease"[Title/Abstract] OR "heart failure"[Title/Abstract] OR "cardiac failure"[Title/Abstract] OR angioplasty OR "percutaneous coronary intervention"[Title/Abstract] OR "cardiovascular mortality"[Title/Abstract] OR mace OR stroke OR angina OR "major adverse cardiovascular event*"[Title/Abstract] OR chd[Title/Abstract] OR acs[Title/Abstract] OR mi[Title/Abstract] OR hf[Title/Abstract] OR pci[Title/Abstract] OR cvd[Title/Abstract] OR sbp[Title/Abstract] OR dpb[Title/Abstract]

#14

#12 OR #13

#15

#1 AND #2 AND #9 AND #14

**OUTCOME: RENAL DISEASE**

#16

((((("Kidney Diseases"[Mesh]) OR "Kidney"[Mesh]) OR "Proteinuria"[Mesh]) OR "Albumins"[Mesh]) OR "Creatinine"[Mesh] OR “glomerular filtration rate”[Mesh])

#17

nephropathy[Title/Abstract] OR kidney*[Title/Abstract] OR proteinuria[Title/Abstract] OR macroalbuminuria[Title/Abstract] OR renal[Title/Abstract] OR ckd[Title/Abstract] OR dkd[Title/Abstract] OR albumin[Title/Abstract] OR eskd[Title/Abstract] OR egfr[Title/Abstract] OR "glomerular filtration rate"[Title/Abstract] OR creatinine[Title/Abstract] OR acr[Title/Abstract]

#18

#16 OR #17

#19

#1 AND #2 AND #9 AND #18

**RCT**

#20

#1 AND #2 AND #8

#### SGLT2i

#21

"Diabetes Mellitus, Type 2"[Mesh] OR TD2[Title/Abstract] OR "Type 2 diabetes"[Title/Abstract] OR "Diabetes, type 2"[Title/Abstract

#22

"Sodium-Glucose Transporter 2 Inhibitors"[Mesh] Or "Sodium-Glucose Transporter 2"[Mesh]

#23

sglt2 OR sglt-2 OR sglt2-inhibitor* OR "sglt2 inhibitor*"[Title/Abstract] OR sglt2inhibitor* OR "sglt-2 inhibitor*"[Title/Abstract] OR sglt2i OR "sodium glucose transporter 2 inhibitor*"[Title/Abstract] OR "sodium glucose transporter-2 inhibitor*"[Title/Abstract] OR dapagliflozin OR empagliflozin OR ertugliflozin OR canagliflozin OR "sodium-glucose transporter 2"[Title/Abstract]

#24

#22 OR #23

#25

White[Title/Abstract] OR Caucasian[Title/Abstract] OR Asian[Title/Abstract] OR African[Title/Abstract] OR Ethnicity[Title/Abstract] OR Ethnic[Title/Abstract]

#26

Age[Title/Abstract] OR "Diabetes duration"[Title/Abstract] OR "BMI"[Title/Abstract] OR "Body Mass Index"[Title/Abstract] OR Adiposity[Title/Abstract] OR "Sex"[Title/Abstract] OR "clinical features"[Title/Abstract]

#27

"Insulin resistant"[Title/Abstract] OR "Insulin deficient"[Title/Abstract] OR "beta-cell function"[Title/Abstract] OR "c-peptide*"[Title/Abstract] OR "metabolite*"[Title/Abstract] OR "Metabolomic"[Title/Abstract] OR "Proteomic"[Title/Abstract] OR "Protein*"[Title/Abstract] OR "peptide*"[Title/Abstract]

#28

(Genotype[Title/Abstract] OR SNP[Title/Abstract] OR variant[Title/Abstract] OR GWAS[Title/Abstract] OR "Genome Wide Association Study"[Title/Abstract] OR Genetic[Title/Abstract] OR Epigenetic[Title/Abstract] OR Methylation[Title/Abstract]) OR ((("Genotype"[Mesh]) OR "Genome-Wide Association Study"[Mesh]) OR "Methylation"[Mesh])

#29

Precision[Title/Abstract] OR Prediction[Title/Abstract] OR Stratification[Title/Abstract] OR Stratified [Title/Abstract] OR Individualised[Title/Abstract] OR Individualisation[Title/Abstract] OR Personalized[Title/Abstract] OR Personalization[Title/Abstract] OR targeted[Title/Abstract]

#30

RCT OR randomis* OR randomiz*

#31

#25 OR #26 OR #27 OR #28 OR #29

**Outcome: HbA1c reduction**

#32

"hba1c reduction"[Title/Abstract] OR "hba1c change*"[Title/Abstract] OR "hba1c response"[Title/Abstract] OR "treatment response"[Title/Abstract] OR "glycaemic efficacy"[Title/Abstract] OR "glycemic efficacy"[Title/Abstract] OR fall in hba1c[Title/Abstract] OR change from baseline in hba1c[Title/Abstract] OR change in hba1c[Title/Abstract] OR changes in hba1c[Title/Abstract] OR slope of change[Title/Abstract] OR "hba1c outcome*"[Title/Abstract] OR outcome hba1c[Title/Abstract] OR "A1C change*"[Title/Abstract] OR "A1C reduction"[Title/Abstract] OR A1C response[Title/Abstract] OR A1C outcome*[Title/Abstract]

#33

#21 AND #24 AND #31 AND #32

**Outcome: Cardiovascular disease**

#34

((((((((("Cardiovascular Diseases"[Mesh]) OR "Myocardial Infarction"[Mesh]) OR "Myocardial Revascularization"[Mesh]) OR "Acute Coronary Syndrome"[Mesh]) OR "Myocardial Ischemia"[Mesh]) OR "Heart Failure"[Mesh]) OR "Angioplasty"[Mesh]) OR "Percutaneous Coronary Intervention"[Mesh]) OR "Stroke"[Mesh]) OR "Angina Pectoris"[Mesh])

#35

"cardiovascular disease"[Title/Abstract] OR "myocardial infarction"[Title/Abstract] OR revasculari*[Title/Abstract] OR "acute coronary syndrome"[Title/Abstract] OR "coronary heart disease"[Title/Abstract] OR "heart failure"[Title/Abstract] OR "cardiac failure"[Title/Abstract] OR angioplasty OR "percutaneous coronary intervention"[Title/Abstract] OR "cardiovascular mortality"[Title/Abstract] OR mace OR stroke OR angina OR "major adverse cardiovascular event*"[Title/Abstract] OR chd[Title/Abstract] OR acs[Title/Abstract] OR mi[Title/Abstract] OR hf[Title/Abstract] OR pci[Title/Abstract] OR cvd[Title/Abstract] OR sbp[Title/Abstract] OR dpb[Title/Abstract]

#36

#34 OR #35

#37

#21 AND #24 AND #31 AND #36

**Outcome: Kidney disease**

#38

((((("Kidney Diseases"[Mesh]) OR "Kidney"[Mesh]) OR "Proteinuria"[Mesh]) OR "Albumins"[Mesh]) OR "Creatinine"[Mesh] OR “glomerular filtration rate”[Mesh])

#39

nephropathy[Title/Abstract] OR kidney*[Title/Abstract] OR proteinuria[Title/Abstract] OR macroalbuminuria[Title/Abstract] OR renal[Title/Abstract] OR ckd[Title/Abstract] OR dkd[Title/Abstract] OR albumin[Title/Abstract] OR eskd[Title/Abstract] OR egfr[Title/Abstract] OR "glomerular filtration rate"[Title/Abstract] OR creatinine[Title/Abstract] OR acr[Title/Abstract]

#40

#38 OR #39

#41

#21 AND #24 AND #31 AND #40

**RCT**

#42

#21 AND #24 AND #30

#### Supplementary Figure 1: Study Screening and Attrition Diagrams

#### a: SGLT2i PRISMA Diagram

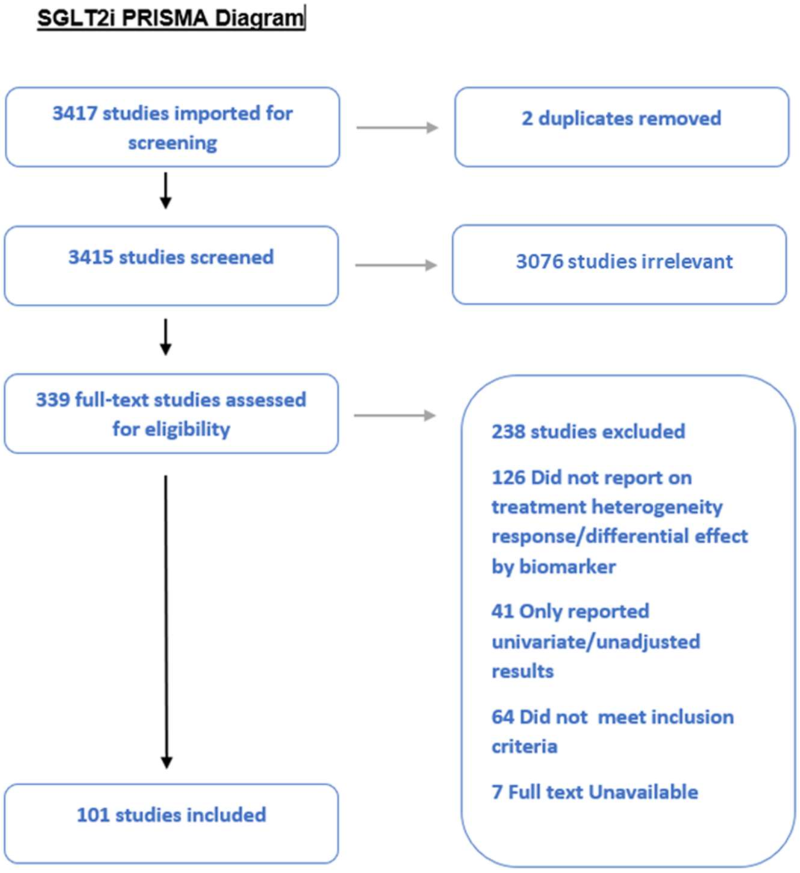

### b: GLP1-RA PRISMA Diagram

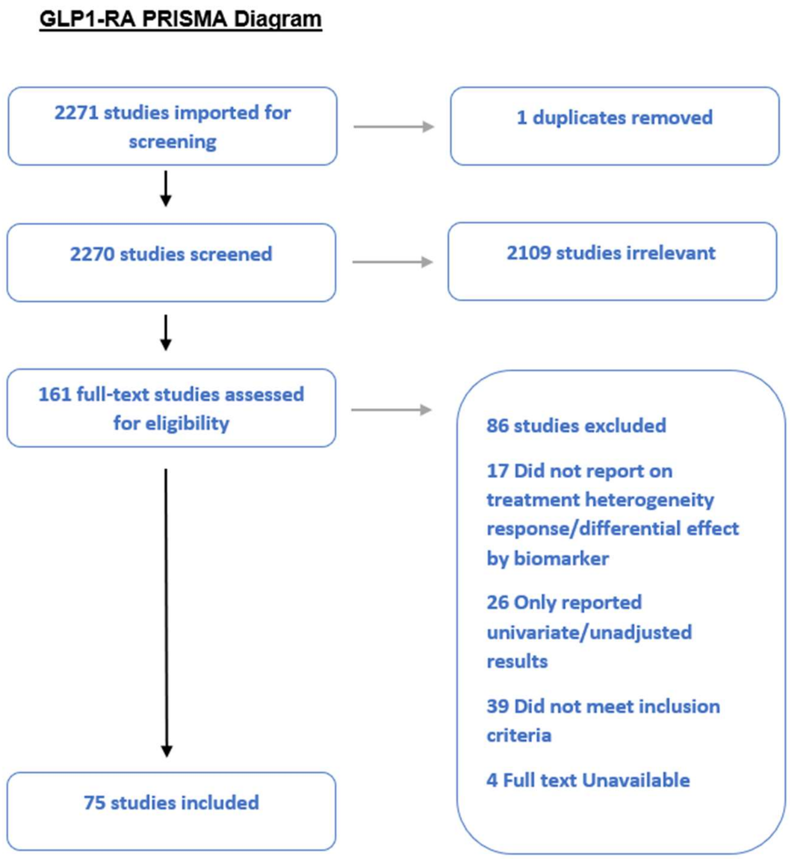

### Supplementary Table: Included studies

#### Supplementary Table 1a SGLT2i and Cardiovascular Disease

| PMID | Author year | Study type | Data sources | Intervention (n) | Comparator (n) | Biomarkers examined | Significant interactions | Notes |
|---|---|---|---|---|---|---|---|---|
| ***Studies examining MACE outcomes*** | | | | | | | | |
| 27896705 | Mahmoud 2017 | Meta-analysis | EMPA-REG, CANVAS | Total = 22256 | | Sex | None | |
| 30786725 | Zelniker 2019 | Meta-analysis | EMPA-REG, CANVAS, DECLARE | Total = 34322 | | CVD history | None | P=0.05 favoring greater SGLT-2 benefit in patients with CVD history |
| 30424892 | Zelniker 2019 | Meta-analysis | EMPA-REG, CANVAS, DECLARE | 19064 | 15258 | Kidney function biomarkers (eGFR, creatinine, UACR); history of CVD; history of heart failure | None | |
| 32993654 | D'Andrea 2020 | Meta-analysis | EMPA-REG, CANVAS, DECLARE | 19064 | 15258 | Age (at 65), sex, race (white, Black, Asian), CVD history, HF history, eGFR (at 60), HbA1c (at 8%), diabetes duration (at 10 years), blood pressure (at 130/80), BMI (at 30) | Greater SGLT benefit in patients with history of CVD | |
| 32165164 | Giugliano 2020 | Meta-analysis | EMPA-REG, CANVAS, DECLARE | Total = 34323 | | Age, statin use | None | |
| 31486272 | Rådholm 2020 | Meta-analysis | EMPA-REG, CANVAS, CREDENCE, DECLARE | NA | NA | Sex | None | |
| 33031522 | McGuire 2021 | Meta-analysis | EMPA-REG, CANVAS, DECLARE, CREDENCE, VERTIS-CV | 26765 | 20204 | CVD history, HF history, eGFR (<60, 60-90, ≥90), ACR (normal, microalbuminuria, macroalbuminuria), HbA1c (<8.5, ≥8.5) | None | |
| 34964748 | Chang 2021 | Meta-analysis | EMPA-REG, CANVAS, DECLARE, CREDENCE, VERTIS-CV, SCORED, SOLOIST-WHF | Total = 5873 | | Age (at 65), sex, race (white, other), diabetes duration (at 10 years), blood pressure (at 140/90), BMI (at 30), diabetes and CVD medication use | None | |
| 34746739 | Chun 2021 | Meta-analysis | EMPA-REG, CANVAS, DECLARE, CREDENCE, VERTIS-CV, DAPA-HF, DAPA-CKD, EMPEROR-R, SCORED, | Total = 61821 | | eGFR (<45, 45-60, ≥60), ACR (normal, microalbuminuria, macroalbuminuria), | Greater SGLT-2 benefit with lower eGFR and higher ACR | |

| PMID | Author year | Study type | Data sources | Intervention (n) | Comparator (n) | Biomarkers examined | Significant interactions | Notes |
|---|---|---|---|---|---|---|---|---|
| | | | SOLOIST-WHF | | | | | |
| 33710721 | Giugliano 2021 | Meta-analysis | EMPA-REG, CANVAS, DECLARE, CREDENCE, VERTIS-CV, SCORED | Total = 65587 | | CVD history | None | |
| 33986377 | Uneda 2021 | Meta-analysis | EMPA-REG, CANVAS, DECLARE, CREDENCE, VERTIS-CV | Total = 102728 | | BMI (at 30) | None | p-value for interaction not given |
| 33707305 | Lee 2021 | Meta-analysis | EMPA-REG, CANVAS, VERTIS-CV | Total = 23556 | | Race (Asian vs. white) | None | |
| 34780865 | Kawai 2022 | Meta-analysis | EMPA-REG, CANVAS, DECLARE, CREDENCE, VERTIS-CV, SCORED | 31646 | 49560 | ACR (normal, microalbuminuria, macroalbuminuria) | None | |
| 34985519 | Bhattarai 2022 | Meta-analysis | EMPA-REG, CANVAS, DECLARE, CREDENCE, VERTIS-CV, CREDENCE DAPA-HF, DAPA-CKD, EMPEROR-R, SCORED, SOLOIST-WHF | 39053 | 32500 | Age (at 65), race (white vs. Asian/Black/other) | None | |
| 28025462 | Kaku 2017 | RCT | EMPA-REG | 4687 | 2333 | Asian vs. other race/ethnicity | None | |
| 29713728 | Zinman 2018 | RCT | EMPA-REG | 4687 | 2513 | Sex | None | |
| 28904068 | Wanner 2018 | RCT | EMPA-REG | 4687 | 2333 | CKD (eGFR <60 and/or ACR ≥300) | None | |
| 29777264 | Verma 2018 | RCT | EMPA-REG | 4687 | 2333 | History of CABG | None | |
| 30882239 | Furtado 2019 | RCT | DECLARE | 8582 | 8578 | CVD history | Greater dapagliflozin benefit with history of CVD | |
| 31530577 | Neuen 2019 | RCT | CANVAS | 5740 | 4293 | ACR (normal, microalbuminuria, macroalbuminuria) | None | |
| 32744354 | Wanner 2020 | RCT | EMPA-REG | 4687 | 2333 | Categories of DKD by ACR and eGFR | None | |
| 32795086 | Bonaca 2020 | RCT | DECLARE | 8582 | 8578 | PAD history | None | |
| 32618884 | Bohm 2020 (#331) | RCT | EMPA-REG | 4687 | 2333 | Decrease in SBP in response to treatment | None | |
| 31820559 | Bohm 2020 (#332) | RCT | EMPA-REG | 4687 | 2333 | AF history | None | |
| 32227432 | Verma 2020 | RCT | EMPA-REG | 4687 | 2333 | TRS-HF-DM score | None | |
| 32485734 | Inzucchi 2020 | RCT | EMPA-REG | 4687 | 2333 | Age, sex, BMI, HbA1c, eGFR, region, CVD risk factor goal attainment | None | |

| PMID | Author year | Study type | Data sources | Intervention (n) | Comparator (n) | Biomarkers examined | Significant interactions | Notes |
|---|---|---|---|---|---|---|---|---|
| 32030863 | Verma 2019 | RCT | EMPA-REG | 4687 | 2333 | Plasma uric acid (tertiles) | None | |
| 33307785 | Langslet 2020 | RCT | EMPA-REG | 4623 | 2309 | LDL cholesterol (5 categories) | None | |
| 33004464 | Neeland 2020 | RCT | EMPA-REG | 4687 | 2333 | History of sleep apnea | None | |
| 33611623 | O'Donoghue 2021 | RCT | DECLARE | 8582 | 8578 | Sex | None | |
| 34427295 | Oyama 2021 | RCT | DECLARE | 8570 | 8564 | BMI, weight, adiposity markers | Greater dapagliflozin benefit in obese vs. non obese | MACE outcome included HHF |
| 33851953 | Zelniker 2021 | RCT | DECLARE | 8582 | 8578 | eGFR, UACR | None | |
| 32971190 | Neuen 2021 | RCT | CANVAS | 5739 | 4292 | KDIGO risk categories | None | |
| 34415356 | Sen 2021 | RCT | CANVAS | 2342 | 1181 | Renal biomarkers: TNFR-1, TNFR-2, KIM-1 | None | |
| 34854308 | Sen 2021 | RCT | CANVAS | 2357 | 1192 | GDF-15 (tertiles) | None | |
| 34908223 | Kaku 2022 | RCT | EMPA-REG | 4623 | 2309 | Asian vs. other race/ethnicity | None | Analyzed total MACE events (rather than time to event) |
| 35115099 | Vaduganathan 2022 | RCT | CANVAS | 2339 | 1164 | Stress cardiac biomarkers: hs-cTnT, sST2, and IGFBP7 | Greater canagliflozin benefit with higher levels of hs-cTnT and sST2, and more elevated biomarkers | |
| 35120199 | Oikonomou 2022 | RCT | CANVAS | 2886 | 1441 | 15-variable algorithm combining demographics and medical history | Algorithm predicted canagliflozin response | |
| 28781064 | Birkeland 2017 | Observational | National registry data in Denmark, Sweden, Norway | 22830 | 68490 | Age, sex, history of renal disease, history of CVD, history of HF, background anti-hyperglycemic therapy, baseline antihypertensive therapy | Qualitative differences: stronger benefit of SGLT2i on MACE seen in older patients and those with pre-existing CVD. | |
| 31902326 | Raparelli 2020 | Observational | Marketscan-Database | 11774 | 89105 | Sex | None | Composite outcome includes HF |
| 34047459 | Chan 2021 | Observational | Chang Gung Memorial Hospital Research Database (Taiwan) | 11769 | na | Age, sex, BMI, weight, adiposity markers, HbA1c | Use of diuretic, prior stroke, older age, female sex, and lower BMI were associated with initial eGFR decline of >30% in those treated with SGLT2i; initial eGFR | |

| PMID | Author year | Study type | Data sources | Intervention (n) | Comparator (n) | Biomarkers examined | Significant interactions | Notes |
|---|---|---|---|---|---|---|---|---|
| | | | | | | | decline of >30% was associated with higher risk of new-onset atrial fibrillation and composite of MACE/HF in SGLT2i-treated individuals. | |
| 33973690 | Idris 2021 | Observational | CPRD Aurum (England) | 24438 | 24438 (DPP4i) | Age, sex, history of renal disease, history of CVD | None | |
| 34570599 | Patorno 2021 | Observational | Medicare and 2 U.S. commercial claims data sets. | 310417 | 246692 | History of CVD | Greater benefit on MACE outcome for SGLT2i compared to GLP1RA in individuals with history of CVD than in those without history of CVD. | See below for HF outcome. |
| 35132865 | Htoo 2022 | Observational | Medicare | 24747 | 22596 | History of CVD, history of heart failure, or both | Relative risk of MACE in SGLT2i-treated compared to GLP1RA-treated was higher in those without history of CVD and without history of HF; similar in those with history of CVD or history of HF; lower in those with history of CVD and history of HF. | See below for HF outcome. |
| ***Studies examining composite heart failure outcomes*** | | | | | | | | |
| 30424892 | Zelniker 2019 | Meta-analysis | EMPA-REG, CANVAS, DECLARE | 19064 | 15258 | | | |
| 31486272 | Rådholm 2020 | Meta-analysis | EMPA-REG, CANVAS, CREDENCE, DECLARE | NA | NA | Sex | None | |
| 33031522 | McGuire 2021 | Meta-analysis | EMPA-REG, CANVAS, DECLARE, CREDENCE, VERTIS-CV | 26765 | 20204 | CVD history, HF history, eGFR (<60, 60-90, ≥90), ACR (normal, microalbuminuria, macroalbuminuria), HbA1c (at 8.5%) | None | |
| 34295696 | Qiu 2021 | Meta-analysis | EMPA-REG, DECLARE, VERTIS-CV, DAPA-HF, EMPEROR-R, SCORED, | NA | NA | Race (white, Black, Asian) CVD history, HF history, CKD history, NYHA HF class, LVEF, geographic region | Greater SGLT benefit in patients with NYHA HF Class II (vs. class III or IV) and in | Subgroup analyses included subsets of CVOTs |

| PMID | Author year | Study type | Data sources | Intervention (n) | Comparator (n) | Biomarkers examined | Significant interactions | Notes |
|---|---|---|---|---|---|---|---|---|
| | | | SOLOIST-WHF | | | | Black and Asian (vs. white) patients | |
| 34964748 | Chang 2021 | Meta-analysis | EMPA-REG, CANVAS, DECLARE, CREDENCE, VERTIS-CV, SCORED, SOLOIST-WHF | Total = 5873 | | Age (at 65), sex, race (white, other), diabetes duration (at 10 years), blood pressure (at 140/90), BMI (at 30), diabetes and CVD medication use | None | |
| 33609071 | Bhatia 2021 | Meta-analysis | EMPA-REG, CANVAS, DECLARE, CREDENCE, VERTIS-CV, DAPA-HF, DAPA-CKD, EMPEROR-R, SCORED, SOLOIST-WHF | Total = 71553 | | Mean age, sex, CVD history, HF history, CKD history, change in mediating biomarkers (HbA1c, blood pressure, body weight) | None | |
| 33707305 | Lee 2021 | Meta-analysis | DAPA-HF, EMPEROR-R | Total = 23556 | | Race (Asian vs. white) | Greater SGLT-2 benefit in Asian vs. white | |
| 34746739 | Chun 2021 | Meta-analysis | EMPA-REG, CANVAS, DECLARE, CREDENCE, VERTIS-CV, DAPA-HF, DAPA-CKD, EMPEROR-R, SCORED, SOLOIST-WHF | Total = 61821 | | eGFR (<45, 45-60, ≥60), ACR (normal, microalbuminuria, macroalbuminuria), | Greater SGLT-2 benefit with lower eGFR and higher ACR | |
| 34985519 | Bhattarai 2022 | Meta-analysis | EMPA-REG, CANVAS, DECLARE, CREDENCE, VERTIS-CV, DAPA-HF, DAPA-CKD, EMPEROR-R, SCORED, SOLOIST-WHF | 39053 | 32500 | Age (at 65), race (white vs. Asian/Black/other) | None | |
| 35020750 | Li 2022 | Meta-analysis | DECLARE, CREDENCE, VERTIS-CV, DAPA-HF, DAPA-CKD, EMPEROR-R, EMPEROR-P, SCORED, SOLOIST-WHF | 14451 | 13372 | eGFR (<30, 30-45, 45-60), HF history, CVD history | None | Population limited to CKD stage 3-4. |
| 26819227 | Fitchett 2016 | RCT | EMPA-REG | 4687 | 2333 | Age (at 65), sex, race, HbA1c (at 8.5%), BMI (at 30), SBP (at 140/90), eGFR (<60, 60-90, ≥90), diabetes and | None | |

| PMID | Author year | Study type | Data sources | Intervention (n) | Comparator (n) | Biomarkers examined | Significant interactions | Notes |
|---|---|---|---|---|---|---|---|---|
| | | | | | | CVD medication use | | |
| 28025462 | Kaku 2017 | RCT | EMPA-REG | 4687 | 2333 | Asian vs. other race/ethnicity | None | |
| 29713728 | Zinman 2018 | RCT | EMPA-REG | 4687 | 2513 | Sex | None | |
| 29526832 | Rådholm 2018 | RCT | CANVAS | 5795 | 4347 | HF history | Greater canagliflozin benefit with history of HF | |
| 28904068 | Wanner 2018 | RCT | EMPA-REG | 4687 | 2333 | CKD (eGFR <60 and/or ACR ≥300) | None | |
| 29777264 | Verma 2018 | RCT | EMPA-REG | 4687 | 2333 | History of CABG | None | |
| 30882239 | Furtado 2019 | RCT | DECLARE | 8582 | 8578 | CVD history | None | |
| 30586757 | Fitchett 2019 | RCT | EMPA-REG | 4687 | 2333 | CVD history, CVD risk score | None | |
| 31530577 | Neuen 2019 | RCT | CANVAS | 5740 | 4293 | ACR (normal, microalbuminuria, macroalbuminuria) | None | |
| 31474116 | Berg 2019 | RCT | DECLARE | 8582 | 8578 | Risk score for HHF | None | |
| 32744354 | Wanner 2020 | RCT | EMPA-REG | 4687 | 2333 | CKD categories by ACR and eGFR | None | |
| 32795086 | Bonaca 2020 | RCT | DECLARE | 8582 | 8578 | PAD history | None | |
| 33004464 | Neeland 2020 | RCT | EMPA-REG | 4687 | 2333 | History of sleep apnea | None | |
| 32030863 | Verma 2019 | RCT | EMPA-REG | 4687 | 2333 | Plasma uric acid (tertiles) | None | |
| 33307785 | Langslet 2020 | RCT | EMPA-REG | 4623 | 2309 | LDL cholesterol (5 categories) | None | |
| 32618884 | Bohm 2020 (#331) | RCT | EMPA-REG | 4687 | 2333 | Decrease in SBP in response to treatment | None | |
| 31820559 | Bohm 2020 (#332) | RCT | EMPA-REG | 4687 | 2333 | AF history | None | |
| 32227432 | Verma 2020 | RCT | EMPA-REG | 4687 | 2333 | TRS-HF-DM score | None | |
| 33611623 | O'Donoghue 2021 | RCT | DECLARE | 8582 | 8578 | Sex | None | |
| 33950573 | Ji 2021 | RCT | EMPA-REG | 4687 | 2333 | 3 BMI categories, stratified by Asian race | None | |
| 34427295 | Oyama 2021 | RCT | DECLARE | 8570 | 8564 | BMI, weight, adiposity markers | Greater dapagliflozin benefit in obese vs. non-obese | |
| 33851953 | Zelniker 2021 | RCT | DECLARE | 8582 | 8578 | eGFR, UACR | None | |
| 32971190 | Neuen 2021 | RCT | CANVAS | 5739 | 4292 | KDIGO risk categories | None | |
| 34364665 | Savarese 2021 | RCT | EMPA-REG | 4675 | 2326 | Cardiac ejection fraction prediction model | None | |
| 34415356 | Sen 2021 | RCT | CANVAS | 2342 | 1181 | Renal biomarkers: TNFR-1, TNFR-2, KIM-1 | None | |
| 34854308 | Sen 2021 | RCT | CANVAS | 2357 | 1192 | GDF-15 (tertiles) | None | |

| PMID | Author year | Study type | Data sources | Intervention (n) | Comparator (n) | Biomarkers examined | Significant interactions | Notes |
|---|---|---|---|---|---|---|---|---|
| 34325887 | Sharma 2021 | RCT | EMPA-REG | Total = 6639 | | Latent class analysis by demographics and clinical features | None | |
| 34908223 | Kaku 2022 | RCT | EMPA-REG | 4623 | 2309 | Asian vs. other race/ethnicity | None | Analyzed total HF events (rather than time to event) |
| 35115099 | Vaduganathan 2022 | RCT | CANVAS | 2339 | 1164 | Stress cardiac biomarkers: hs-cTnT, sST2, and IGFBP7 | None | |
| 29935543 | Kim 2018 | Observational | Korean Health Insurance Review and Assessment Service | 59479 | 59479 (DPP4i) | History of CVD | Relative benefit in reducing HF hospitalization with SGLT2i vs DPP4i treatment seen more quickly in those with CVD history than in those without | |
| 31902326 | Raparelli 2020 | Observational | Marketscan-Database | 11774 | 89105 (SU) | Sex | None | Composite outcome includes HF |
| 33599357 | Becher 2021 | Observational | Swedish heart failure registry | 361 | 1083 | Kidney function biomarkers (eGFR, creatinine, UACR), background anti-hyperglycemic therapy, heart failure type (HFpEF, HFmrEF, HFrEF) | No interactions observed by HF subtype, concomitant metformin treatment, or eGFR | |
| 33973690 | Idris 2021 | Observational | CPRD Aurum (England) | 24438 | 24438 (DPP4i) | Age, sex, history of renal disease, history of CVD | None | |
| 34570599 | Patorno 2021 | Observational | Medicare and 2 U.S. commercial claims data sets. | 310417 (SGLT2i) | 246692 (GLP1RA) | History of CVD | No difference in relative benefit of SGLT2i compared to GLP1RA in individuals with or without history of CVD. | See above for MACE outcome. |
| 35132865 | Htoo 2022 | Observational | Medicare | 24747 (SGLT2i) | 22596 (GLP1RA) | History of CVD, history of heart failure, or both | Relative reduction in HF hospitalization was greater for SGLT2i compared to GLP1RA in those with history of HF but not of CVD. | See above for MACE outcome. |

#### Supplementary table 1b: GLP1RA and Cardiovascular Outcomes

| PMID | Author (year) | Study type | Data sources | Intervention (n) | Comparator (n) | Biomarkers examined | Significant interactions | Notes |
|---|---|---|---|---|---|---|---|---|
| 30566004 | Verma 2018 | RCT | LEADER | 4668 | 4672 | History of CVD | Liraglutide reduced risk of CV outcomes in individuals with history of MI/stroke, but not in those without history. | |
| 32744418 | Verma 2020 | RCT | LEADER; SUSTAIN 6 | 6316 | 6321 | Adiposity biomarkers | Semaglutide more efficacious with higher baseline BMI. | Only seen in SUSTAIN 6, LEADER showed no GLP1RA/biomarker interaction. |
| 30851070 | Verma 2019 | RCT | LEADER; SUSTAIN 6 | NA | NA | Diabetes duration | None | |
| 32643857 | Verma 2020 | RCT | LEADER; SUSTAIN 6 | NA | NA | Microvascular disease | None | |
| 33537745 | Riddle 2021 | RCT | REWIND | 4949 | 4952 | Age | None | |
| 32618386 | Mosenzon O 2020 | RCT | LEADER | 4668 | 4672 | UACR and eGFR strata | None | |
| 30371301 | Mentz 2018 | RCT | EXSCEL | 7356 | 7356 | Cardiovascular risk score | None | |
| 32164886 | Marso 2020 | RCT | LEADER | 4668 | 4672 | History of heart failiure | None | |
| 30566006 | Mann 2018 | RCT | LEADER | 4668 | 4672 | Kidney function biomarkers | None | |
| 31167654 | Leiter 2019 | RCT | SUSTAIN 6 | 1648 | 1649 | Age; Sex; History of CVD | None | |
| 32372454 | Leiter 2020 | RCT | LEADER; SUSTAIN 6 | NA | NA | Blood pressure strata | None | |
| 30508430 | Gilbert 2019 | RCT | LEADER | 4668 | 4672 | Age | None | |
| 31542942 | Fudim 2019 | RCT | EXSCEL | 7356 | 7396 | History of heart failiure | Exenatide had attenuated efficacy in individuals with HF at baseline. | |
| 33905751 | Barbery 2021 | RCT | EXSCEL | 7356 | 7396 | Sex; Ethnicity; History of CVD; History of heart failiure; Smoking status | Individuals on exenatide enrolled in Latin America experienced lower risk of ACS or coronary revascularisation. Exenatide had no differential effect in individuals with or without CVD history. | |
| 31752517 | Badjatiya 2019 | RCT | EXSCEL | 7355 | 7396 | History of CVD | None | Exenatide had no effect on CVD risk in individuals with or without PAD. |
| 29935211 | Wang 2018 | Meta-analysis | LEADER; ELIXA; | 16706 | 16751 | Sex; Ethnicity; Adiposity | CV benefits associated with | |

| PMID | Author (year) | Study type | Data sources | Intervention (n) | Comparator (n) | Biomarkers examined | Significant interactions | Notes |
|---|---|---|---|---|---|---|---|---|
| | | | SUSTAIN 6; EXSCEL; | | | biomarkers; Kidney function biomarkers; History of CVD; Background anti-hyperglycaemic therapy | GLP-1RA use was only observed in male, black, Asian, or obese patients. | |
| 34144086 | Tsapas 2021 | Meta-analysis | HARMONY OUTCOMES; REWIND; EXSCEL; LEADER | NA | NA | Background treatment with metformin | None | |
| 32077924 | Marsico 2020 | Meta-analysis | ELIXA; LEADER; SUSTAIN 6; EXSCEL; HARMONY; REWIND; PIONEER 6 | 27977 | 28027 | History of CVD with or without prior MI; History of CVD; Cardiovascular risk factors | None | |
| 31903692 | Husain 2020 | Pooled RCT | SUSTAIN 6; PIONEER 6 | 3239 | 3241 | History of CVD; History of CV outcome | Semaglutide less efficacious in individuals with prior HF. | Semaglutide showed consistent effects on MACE versus comparators across CV risk categories. |
| 32998732 | Husain 2020 | Pooled RCT | SUSTAIN 6; PIONEER 6 | 10508 | 7137 | Cardiovascular risk score | Relative risk reduction was largest in the low CV risk score group; the largest absolute risk reduction was in the intermediate/high CV risk score group. | |
| 32734559 | He 2020 | Meta-analysis | LEADER; SUSTAIN 6; EXSCEL; HARMONY OUTCOMES; REWIND; PIONEER 6 | 24583 | 24633 | Age; Sex; Diabetes duration; Adiposity biomarkers; Glycaemic biomarkers; Kidney function biomarkers; History of CVD; Geographic location | None | Slight trend observed suggesting established CVD may have increased benefit to GLP-1RAs. |
| 31373167 | Giugliano 2019 | Meta-analysis | REWIND; PIONEER 6 | NA | NA | History of CVD | None | |
| 34780865 | Kawai 2022 | Meta-analysis | ELIXA; LEADER; REWIND | 24362 | 15936 | Kidney function biomarkers; Albuminuria status | None | |
| 31595657 | Mannucci 2019 | Meta-analysis | ELIXA; LEADER; SUSTAIN 6; EXSCEL; HARMONY; REWIND; PIONEER-6 | 27977 | 28027 | Age; Sex; Adiposity biomarkers; Kidney function biomarkers; History of CVD; | None | |

| PMID | Author (year) | Study type | Data sources | Intervention (n) | Comparator (n) | Biomarkers examined | Significant interactions | Notes |
|---|---|---|---|---|---|---|---|---|
| | | | | | | Geographic location | | |
| 30786725 | Zelniker 2019 | Meta-analysis | ELIXA; LEADER; SUSTAIN-6; EXSCEL; HARMONY; EMPA-REG OUTCOME | NA | NA | History of cardiovascular disease | GLP1RAs only reduced MACE in patients with established CVD compared to those without; | |
| 33986377 | Uneda 2021 | Meta-analysis | ELIXA; EXSCEL; LEADER; HARMONY Outcomes; PIONEER 6; REWIND; SUSTAIN 6 | NA | NA | Adiposity biomarkers | None | |
| 32142999 | Singh 2020 | Meta-analysis | ELIXA; LEADER; SUSTAIN 6; EXSCEL; HARMONY; REWIND; PIONEER 6 | NA | NA | Sex | None | |
| NA | Qiu 2020 | Meta-analysis | ELIXA; LEADER; SUSTAIN-6; EXSCEL; HARMONY OUTCOMES; REWIND; PIONEER 6 | NA | NA | Sex; Diabetes duration; History of CVD; Background anti-hyperglycaemic therapy | None | |
| 33707305 | Lee 2021 | Meta-analysis | LEADER; SUSTAIN 6; EXSCEL; HARMONY OUTCOMES; REWIND; PIONEER 6 | NA | NA | Ethnicity | GLP1RA use in Asians with T2D associated with reduced CVD outcome risk. | |
| 32993654 | D'Andrea 2020 | Meta-analysis | ELIXA; LEADER; SUSTAIN 6; EXSCEL; HARMONY; REWIND; PIONEER 6 | 27681 | 27757 | Age; Sex; Diabetes duration; Ethnicity; Adiposity biomarkers; Glycaemic biomarkers; Kidney function biomarkers; History of CVD; History of heart failiure; Blood pressure | GLP1RAs only showed CVD protective effect in individuals with prior CVD. | Trend towards greater cardiovascular protection in individuals with uncontrolled diabetes. Uncontrolled hypertension, obesity, gender, age, and race did not modify the effect of GLP1RAs. |
| 32534570 | Yang 2020 | Observational | National Health Insurance Research Database of Taiwan | 5089 | 5089 | Age; Sex; Diabetes duration; History of CVD; Microvascular disease | GLP1RA versus DPP-4i yielded greater cardiovascular benefit in those without established CVD versus those with established CVD. | |
| 35254430 | Chen 2022 | Observational | National Health Insurance | 701 | 26578 | Age; Sex; Cardiovascular outcome; ACE | Lower risk of mortality associated with | |

| PMID | Author (year) | Study type | Data sources | Intervention (n) | Comparator (n) | Biomarkers examined | Significant interactions | Notes |
|---|---|---|---|---|---|---|---|---|
| | | | Research Database of Taiwan | | | inhibitor use; End stage kidney disease status | use of GLP1RAs compared with DDP-4 inhibitors among patients with cerebrovascular disease than those without. | |
| 34570599 | Patorno 2021 | Observational | Medicare and 2 U.S. commercial claims data sets. | 246692 | 310417 | History of CVD | Greater cardiovascular benefit in individuals with CVD compared to without. | |
| 31902326 | Raparelli 2020 | Observational | Marketscan-Database | 14697 | 152557 | Sex | Greater cardiovascular effectiveness in women. | |
| 35132865 | Htoo 2022 | Observational | Medicare | 22596 | 24747 | History of CVD; History of Cardiovascular Outcome | Atherosclerotic CVD events were less frequent with GLP1RA in those without prior CVD or HF. | |

#### Supplementary Table 1c: SGLT2i and renal disease

| PMID | Author year | Study type | Data sources/trials | N (intervention) | N (comparator) | Biomarkers examined | Significant interactions | Notes |
|---|---|---|---|---|---|---|---|---|
| ***Studies examining eGFR changes / CKD progression / composite outcomes of these with or without ACR changes*** | | | | | | | | |
| 31506585 | Bae 2019 | Meta-analysis | 48 studies | 34,661 | 23,504 | eGFR (continuous) | None | Non-significant greater SGLT-2 benefit with higher eGFR |
| 34964748 | Chang 2021 | Meta-analysis | EMPA-REG, CANVAS, DECLARE, CREDENCE (VERTIS-CV, SCORED, SOLOIST-WHF not used for renal outcomes) | (58,783 in intervention + comparator arm, not given separately) | (58,783 in intervention + comparator arm, not given separately) | Age (<65, ≥65 years), sex, race (white, other), diabetes duration (<10, ≥10 years), blood pressure (SBP≥140 or DBP≥90, SBP<140 and DBP<90 mmHg) , BMI (<30, ≥30 kg/m$^2$), diabetes and CVD medication use | None | |
| 34746739 | Chun 2021 | Meta-analysis | EMPA-REG, CANVAS, DECLARE, CREDENCE, DAPA-CKD, SCORED (DAPA-HF, VERTIS-CV, EMPEROR-R, SOLOIST-WHF not used for | (61,821 in intervention + comparator arm, not given separately) | (61,821 in intervention + comparator arm, not given separately) | eGFR (<45, 45-<60, ≥60), ACR (<30 [normoalbuminuria], 30-300 [microalbuminuria], >300 [macroalbuminuria]) | None | |

| PMID | Author year | Study type | Data sources/trials | N (intervention) | N (comparator) | Biomarkers examined | Significant interactions | Notes |
|---|---|---|---|---|---|---|---|---|
| | | | renal outcomes) | | | | | |
| 33710721 | Giugliano 2021 | Meta-analysis | EMPA-REG, CANVAS, DECLARE, CREDENCE, VERTIS-CV, SCORED, DAPA-CKD, EMPEROR-R | (65,587 in intervention + comparator arm, not given separately) | (65,587 in intervention + comparator arm, not given separately) | CVD history | None | |
| 33031522 | McGuire 2021 | Meta-analysis | EMPA-REG, CANVAS, DECLARE, CREDENCE, VERTIS-CV | (46,969 in intervention + comparator arm, not given separately) | (46,969 in intervention + comparator arm, not given separately) | CVD history, HF history, ACR (<30 [normoalbuminuria], 30-300 [microalbuminuria], >300 [macroalbuminuria]) | None | |
| 31495651 | Neuen 2019 (SGLT2 inhibitors for the prevention of kidney failure in patients with type 2 diabetes: a systematic review and meta-analysis) | Meta-analysis | EMPA-REG, CANVAS, DECLARE, CREDENCE | (38,723 in intervention + comparator arm, not given separately) | (38,723 in intervention + comparator arm, not given separately) | CVD medication (RAS blockade) use, eGFR (<45, 45-<60, 60-<90, ≥90), ACR (<30 [normoalbuminuria], 30-300 [microalbuminuria], >300 [macroalbuminuria]) | None | Non-significant trend for greater SGLT2i effect in those with higher baseline eGFR |
| 30424892 | Zelniker 2019 | Meta-analysis | EMPA-REG, CANVAS, DECLARE | 19,064 | 15,258 | CVD status (ASCVD vs multiple risk factors), eGFR (<60, 60-<90, ≥90) | None for CVD status; greater SGLT-2 benefit in patients with higher eGFR | |
| 33214158 | Bakris 2020 | RCT | CREDENCE | 2,202 | 2,199 | eGFR (<30, ≥30) | None | |
| 31820559 | Bohm 2020 (Efficacy of empagliflozin on heart failure and renal outcomes in patients with atrial fibrillation: data from the EMPA-REG OUTCOME trial) | RCT | EMPA-REG | 4,687 | 2,333 | AF history | None | |
| 32795086 | Bonaca 2020 | RCT | DECLARE | 8,582 | 8,578 | PAD history | None | |
| 33158949 | Januzzi 2021 | RCT | CANVAS | 2,384 | 1,193 | IGFBP7 (quartiles) | None | |
| 33619120 | Jardine 2021 | RCT | CREDENCE | 2,202 | 2,199 | ACR (all in macroalbuminuria range: ≤1000, >1000-<3000, ≥3000) | None | |
| 33950573 | Ji 2021 | RCT | EMPA-REG | 4,687 | 2,333 | 3 BMI categories (different for Asian vs non-Asian), stratified by Asian race | None | |
| 30412655 | Kadowaki 2019 | RCT | EMPA-REG | 4,687 | 2,333 | Race (Asian, other) | None | |
| 34233928 | Mosenzon 2021 | RCT | DECLARE | 8,582 | 8,578 | ACR (≤15, 15<30, 30-300 [microalbuminuria], | None for renal outcome; greater SGLT-2 benefit | |

| PMID | Author year | Study type | Data sources/trials | N (intervention) | N (comparator) | Biomarkers examined | Significant interactions | Notes |
|---|---|---|---|---|---|---|---|---|
| | | | | | | >300 [macroalbuminuria]) | with higher ACR for cardiorenal outcome (micro- and macroalbuminuria have greater benefit than other categories) | |
| 33004464 | Neeland 2020 | RCT | EMPA-REG | 4,687 | 2,333 | History of sleep apnea | None | Very small numbers with sleep apnea |
| 31530577 | Neuen 2019 (Effect of Canagliflozin on Renal and Cardiovascular Outcomes across Different Levels of Albuminuria: Data from the CANVAS Program) | RCT | CANVAS | 5,740 | 4,293 | ACR (<30 [normoalbuminuria], 30-300 [microalbuminuria], >300 [macroalbuminuria]) | Greater SGLT-2 benefit with higher ACR for preventing eGFR decline. Also heterogeneity in composite renal outcome by baseline UACR (no benefit of SGLT-2 in microalbuminuria but large benefit for normo- or macroalbuminuria) | |
| 32971190 | Neuen 2021 | RCT | CANVAS | (10,031 in intervention + comparator arm, not given separately) | (10,031 in intervention + comparator arm, not given separately) | KDIGO risk categories (composite of eGFR and ACR) | None | |
| 33611623 | O'Donoghue 2021 | RCT | DECLARE | 8,582 | 8,578 | Sex | None | |
| 34427295 | Oyama 2021 | RCT | DECLARE | 8,570 | 8,564 | BMI (18.5 to <25, 25 to <30, 30 to <35, 35 to <40, ≥40 kg/m$^2$) | None | |
| 34854308 | Sen 2021 (Association Between Circulating GDF-15 and Cardio-Renal Outcomes and Effect of Canagliflozin: Results from the CANVAS Trial) | RCT | CANVAS | 2,357 | 1,192 | GDF-15 (tertiles) | None | |
| 34415356 | Sen 2021 (Effects of the SGLT2 inhibitor canagliflozin on plasma biomarkers TNFR-1, TNFR-2 and KIM-1 in the CANVAS trial) | RCT | CANVAS | 2,342 | 1,181 | Renal biomarkers: TNFR-1, TNFR-2, KIM-1 (tertiles and continuous) | None | Greater SGLT-2 benefit in lowest TNFR-2 tertile (although few events), but non-significant when TNFR-2 treated as a continuous variable |

| PMID | Author year | Study type | Data sources/trials | N (intervention) | N (comparator) | Biomarkers examined | Significant interactions | Notes |
|---|---|---|---|---|---|---|---|---|
| 35115099 | Vaduganathan 2022 | RCT | CANVAS | 2,339 (fewer in total for sST2 and more for IGFBP7) | 1,164 (fewer in total for sST2 and more for IGFBP7) | Stress cardiac biomarkers: hs-cTnT (<14, ≥14 pg/mL and continuous), sST2 (<35, ≥35 ng/mL and continuous), and IGFBP7 (<96.5, ≥96.5 ng/mL and continuous) | None | |
| 29777264 | Verma 2018 | RCT | EMPA-REG | 4,687 | 2,333 | History of CABG | None | |
| 32030863 | Verma 2020 | RCT | EMPA-REG | 4,686 | 2,326 | Plasma uric acid (tertiles) | None | |
| 32744354 | Wanner 2020 | RCT | EMPA-REG | 4,687 | 2,333 | CKD (composite of eGFR <60 and/or ACR ≥300) | None | |
| 33118320 | Koh 2021 | Observational | CVD-REAL 3 Korea (insurance database) | 45,016 | 45,016 | Age (<65, ≥65 years), sex, BMI (<25, ≥25 kg/m$^2$), abdominal obesity, diabetic retinopathy, hypertension, CVD history, eGFR (<60, 60-<90, ≥90), proteinuria, eGFR + proteinurea(<60 + absent, ≥60 + absent, <60 + present, ≥60 + present) | Greater SGLT-2 benefit with higher BMI, with abdominal obesity, and with lower eGFR (<90 vs ≥90) | |
| 34593566 | Nagasu 2021 | Observational | Japan Chronic Kidney Disease Database (registry) | 1,033 | 1,033 | Age (<65, ≥65 years), eGFR (<60, ≥60), rapid decline in eGFR before initiating treatment, proteinuria, ACEi/ARB use | Greater SGLT-2 benefit on eGFR when no rapid eGFR decline and when ACEi/ARB not used | No interaction for composite renal outcome of eGFR decline and ESRD |
| ***Studies examining ACR changes*** | | | | | | | | |
| 31506585 | Bae 2019 | Meta-analysis | 48 studies | 34,661 | 23,504 | ACR (continuous) | None | Non-significant greater SGLT-2 benefit with higher ACR |
| 33158949 | Januzzi 2021 | RCT | CANVAS | 2,384 | 1,193 | IGFBP7 (quartiles) | Greater SGLT-2 benefit on 'first progression of albuminuria' outcome with higher ICFBP7 | |
| 30412655 | Kadowaki 2019 | RCT | EMPA-REG | 4,687 | 2,333 | Race (Asian, other) | None | |

Abbreviations: high-sensitivity cardiac troponin T (hs-cTnT), soluble suppression of tumorigenesis-2 (sST2), insulin-like growth factor binding protein 7 (IGFBP7), estimated glomerular filtration rate in mL/min/1.73m$^2$ (eGFR), albumin/creatinine ratio in mg/g creatinine (ACR), tumor necrosis factor receptor 1 (TNFR-1), tumor necrosis factor receptor 2 (TNFR-2), kidney injury molecule-1 (KIM-1), growth differentiation factor-15 (GDF-15), Kidney Disease Improving Global Outcomes (KDIGO), coronary artery bypass grafting (CABG), peripheral arterial disease (PAD), atrial fibrillation (AF), renin–angiotensin system (RAS)

Studies not included in the Supplementary table 1b above

| Author year | Reason for not including |
|---|---|
| Bohm 2020 (Heart failure and renal outcomes according to baseline and achieved blood pressure in patients with type 2 diabetes: results from EMPA-REG OUTCOME) | Did not report statistical test of interaction |
| Chan 2021 | No control group |
| Idris 2021 | Did not report statistical test of interaction |
| Kawai 2022 | Did not report statistical test of interaction |
| Lin 2021 | Did not report statistical test of interaction |
| Nunoi 2019 | Did not report statistical test of interaction |
| Xie 2020 | Did not report statistical test of interaction |
| Zhou 2019 | Machine learning model identified combinations of biomarkers rather than individual ones |

Supplementary table 1d: GLP1RA and renal disease

| PMID | Author year | Study type | Data sources/trials | Intervention (n) | Comparator (n) | Biomarkers examined | Significant interactions | Notes |
|---|---|---|---|---|---|---|---|---|
| ***Studies examining eGFR changes / CKD progression / composite outcomes of these with or without ACR changes*** | | | | | | | | |
| 32372454 | Leiter 2020 | RCT | LEADER, SUSTAIN 6 (analysed separately) | (9,340 [LEADER], 3,297 [SUSTAIN 6] in intervention + comparator arm, not given separately) | (9,340 [LEADER], 3,297 [SUSTAIN 6] in intervention + comparator arm, not given separately) | Blood pressure (<120/80 [normal], SBP 120-129 and DBP<80 [elevated], SBP 130-139 or DBP 80-89 [stage 1 hypertension], SBP≥140 or DBP≥90 mmHg [stage 2 hypertension]) | None | |
| 32164886 | Marso 2020 | RCT | LEADER | 4,668 | 4,672 | HF history | None | |
| 32618386 | Mosenzon 2020 | RCT | LEADER | 4,668 | 4,672 | eGFR (<30, 30-<45, 45-<60, 60-<90, ≥90), ACR (0, >0-<15, 15-<30, 30-<100, 100-<300 [microalbuminuria], ≥300 [macroalbuminuria]) | None | |
| 30292589 | Muskiet 2018 | RCT | ELIXA | 2,984 | 2,994 | ACR (<30 [normoalbuminuria], 30-300 [microalbuminuria], >300 [macroalbuminuria]) | None | |
| 33537745 | Riddle 2021 | RCT | REWIND | 4,949 | 4,952 | Age (at 65) | None | |
| 34903039 | Shaman 2022 | Pooled RCT | LEADER, SUSTAIN 6 | 6,316 | 6,321 | eGFR (<30, 30-<60, 60-<90, ≥90) | Greater GLP1RA benefit with eGFR 30-60 (other categories: no treatment effect) | |
| 32803900 | vanderAart-vanderBeek 2020 | RCT | EXSCEL | 6,906 | 6,920 | Blood pressure (SBP≥140, SBP<140 mmHg), BMI (<30, ≥30 kg/m²), eGFR (<60, ≥60), CVD history, CVD medication (RAASi) | Greater GLP1RA benefit with lower BMI | Also looked at ACR (≤30, >30-100, >100-200, >200): greater GLP1RA benefit on eGFR decline with higher ACR although no statistical test for interaction |

| PMID | Author year | Study type | Data sources/trials | Intervention (n) | Comparator (n) | Biomarkers examined | Significant interactions | Notes |
|---|---|---|---|---|---|---|---|---|
| 30851070 | Verma 2019 | RCT | LEADER, SUSTAIN 6 (analysed separately) | (9,321 [LEADER], 3,297 [SUSTAIN 6] in intervention + comparator arm, not given separately) | (9,321 [LEADER], 3,297 [SUSTAIN 6] in intervention + comparator arm, not given separately) | Diabetes duration (<5, 5-<15, 15-<25, ≥25 years) | None | |
| 32744418 | Verma 2020 | RCT | LEADER, SUSTAIN 6 (analysed separately) | 4,668 (LEADER), 1,648 (SUSTAIN 6) | 4,672 (LEADER), 1,649 (SUSTAIN 6) | BMI (<25, 25-<30, 30-<35, ≥35 kg/m²) | None | |
| ***Studies examining ACR changes*** | | | | | | | | |
| 30292589 | Muskiet 2018 | RCT | ELIXA | 2,984 | 2,994 | ACR (<30 [normoalbuminuria], 30-300 [microalbuminuria], >300 [macroalbuminuria]) | Greater GLP1 benefit with higher ACR (no treatment effect with normoalbuminuria but strong reduction in proteinuria in those with micro and macroalbumununria) | |
| 34903039 | Shaman 2022 | Pooled RCT | LEADER, SUSTAIN 6 | 6,316 | 6,321 | ACR (<30 [normoalbuminuria], 30-300 [microalbuminuria], >300 [macroalbuminuria]) | Greater GLP1 benefit with microalbuminuria than normo- or macroalbuminuria | |
| 32803900 | vanderAart-vanderBeek 2020 | RCT | EXSCEL | 6,906 | 6,920 | Blood pressure (SBP≥140, SBP<140 mmHg), BMI (<30, ≥30 kg/m²), eGFR (<60, ≥60), CVD history, CVD medication (RAASi) | None | |

Abbreviations: estimated glomerular filtration rate in mL/min/1.73m² (eGFR), albumin/creatinine ratio in mg/g creatinine (ACR), renin-angiotensin-aldosterone system inhibitors (RAASi)

#### Studies not included in the table above

| Author year | Reason for not including |
|---|---|
| Kawai 2022 | Did not report statistical test of interaction |
| Xie 2020 | Did not report statistical test of interaction |

#### Supplementary 1 e. SGLT2i and glycaemic outcomes

| PMID | Author (year) | Study type | Data Source | Intervention (n) | Comparator (n) | Biomarkers examined | Significant interactions | Notes |
|---|---|---|---|---|---|---|---|---|
| 32821142 | Scheen (2020) | Meta-Analysis | 7 RCTs of Asian patients; 16 RCTs of Non-Asian patients | 1164 | 1088 | Ethnicity | None | Greater reduction in HbA1c for Asians vs Non-Asians |
| 29029369 | Cai (2018) | Meta-Analysis | 17 RCTs of Asian patients; 39 RCTs of Non-Asian patients | 5679 | 4170 | Ethnicity | None | |
| 35314533 | Wang (2022) | Meta-Analysis | Adhimadhyam (2018); Amos (2016); Cefalu (2015); Chilton: cohort 1 (2015); Chilton: cohort2 | 287 | 921 | Age | None | SGLT2i benefitted patients aged |

| | | | | | | | | |
|---|---|---|---|---|---|---|---|---|
| | | | (2015); Gautam (2017); Johnson (2017); Kobayashi (2019); Kohler (2016); Leiter (2014); Maegawa (2018); Osonoi (2018); Shiba (2017); Sinclair (2014) | | | | | <65 years for HbA1c reduction |
| 28583425 | Wilding (2017) | Observational | | 5825 | 0 | Background AHA | None | |
| 27822077 | Scheerer (2016) | Observational | | 1169 | 0 | Age; Sex; History of CVD; Background AHA | Higher HbA1c at baseline was significantly associated with greater HbA1c reduction | |
| 32744394 | Montvida (2020) | Observational | Centricity Electronic Medical Records Database (USA) | 82694 | 0 | Ethnicity | Black patients less likely to achieve glycaemic control (HbA1c < 7.5% or 1% reduction) at 18 months than white patients | White and black patients had similar HbA1c reduction at 6 months |
| 31369642 | Cho (2019) | Observational | Korean outpatient clinic | 374 | 0 | Age; Sex; Diabetes duration; Adiposity markers; Glycaemic biomarker | Hba1c reductions greater in younger patients (≤50 years) compared to older patients (>60 years) | HbA1c reduction significantly greater for patients with higher baseline HbA1c; no difference in glycaemic efficacy for BMI subgroups |
| 33240585 | Chen (2020) | Observational | Multicenter observational patient data (Taiwan) | 1197 | 0 | Age; Sex; Adiposity markers; Glycaemic biomarker; Background AHA | Patients with higher baseline HbA1c showed greater reduction in HbA1c at 6 months | |
| 28829163 | Brown (2017) | Observational | Canadian diabetes registry | 1520 | 0 | Age; Sex; Diabetes duration; Adiposity markers; Glycaemic biomarker; History of Renal Disease; CV biomarkers; History of hypertension | Greater reductions in HbA1c seen in patients taking dapagliflozin with higher baseline HbA1c, shorter duration of diabetes, or male sex | |

| | | | | | | | | |
|---|---|---|---|---|---|---|---|---|
| 32424798 | Strain (2020) | Observational | | 490 | 6680 | Background AHA | SGLT2is associated with greater HbA1c reduction in younger patients | Males, lower baseline BMI, and higher baseline DBP also associated with better response for SGLT2is (NS) |
| 30688052 | Lee (2019) | Observational | Korean outpatient clinic | 804 | 0 | Adiposity markers; Background AHA | Higher HbA1c at baseline was significantly associated with greater HbA1c reduction | |
| 31050099 | Zhou (2019) | Observational | Medical Data Vision Database (Japan) | 990 | 4257 | Diabetes duration; Kidney function biomarkers; CV biomarkers; Glycaemic biomarkers; Use of anti-thrombotic agents | None | |
| 30815552 | DeFronzo (2017) | Pooled RCT | EMPA-REG MONO; EMPA-REG H2H-SU | 1213 | 1003 | Glycaemic markers | Patients with higher baseline HbA1c on empagliflozin showed significantly greater reductions in HbA1c than those on sitagliptin or glimepiride | |
| 28860019 | Cherney (2018) | Pooled RCT | Haring (2013); Haring (2014); Kovacs (2014); Roden (2013); Barnett (2014) | 1142 | 1144 | Kidney function markers | None | HbA1c reduction greater for higher baseline eGFR subgroups (≥60 ml/min/1.72m^2) |
| 24742013 | Sinclair (2014) | Pooled RCT | Stenlof (2013); Lavalle-Gonzalez (2013); Wilding (2013) | 1667 | 646 | Age | None | |
| 25059406 | Yamout (2014) | Pooled RCT | Stenlof (2014); Yale (2013); Bode (2013); Neal (2013) | 703 | 382 | Kidney function markers | None | LS mean reductions in HbA1c for patients with stage 3a CKD was greater than those with stage 3b CKD; no formal statistical |

| | | | | | | | | |
|---|---|---|---|---|---|---|---|---|
| | | | | | | | | testing performed performed |
| 26600115 | Matthews (2016) | Pooled RCT | Stenlof (2013); Lavalle-Gonzalez (2013); Wilding (2013); Forst (2014); Schernthaner (2013) | 1316 | 1156 | Glycaemic markers; HOMA2-%B and HOMA2-%S markers | None | Investigated baseline beta-cell function and insulin sensitivity via HOMA2-%B/S tertiles |
| 32324082 | Liu (2020) | Pooled RCT | | 1029 | 515 | Ethnicity | None | |
| 27052454 | Gilbert (2016) | Pooled RCT | Stenlof (2013); Lavalle-Gonzalez (2013); Wilding (2013); Forst (2013); Neal (2015); Fulcher (2015) | 2763 | 1176 | Age; Sex; Adiposity markers; Kidney function markers | None | |
| 26373629 | Blonde (2015) | Pooled RCT | NCT0108183 NCT01106677NCT01106625NCT01106690 | 1667 | 646 | Age; Sex | None | |
| 31933292 | Parigi (2019) | Pooled RCT | Lewin (2015); DeFronzo (2015) | 1080 | 261 | Age; Sex; Diabetes duration; Ethnicity; Adiposity markers; Glycaemic markers; Kidney function biomarkers | None | |
| 32700421 | Pratley (2020) | Pooled RCT | VERTIS-MONO; VERTIS-MET; VERTIS-SITA2; VERTIS-SU; VERTIS-SITA; VERTIS-FACTORIAL;VERTIS-RENAL | 2533 | 1072 | Age | None | No formal statistical testing performed |
| 33004464 | Neeland (2020) | RCT | EMPA-REG OUTCOME | 4687 | 2333 | Obstructive Sleep Apnea | None | |
| 29573139 | Frias (2018) | RCT | DURATION-8 | 233 | 462 | Age; Sex; Diabetes duration; Ethnicity; Adiposity markers; Glycaemic markers; Kidney function biomarkers | None | NS association for age and baseline eGFR; dapagliflozin + exenatide resulted in larger HbA1c reduction than either AHA alone |
| 35233908 | Cherney (2022) | RCT | VERTIS-CV | 5499 | 2747 | Kidney function biomarkers | HbA1c reductions lowest in patients with high/very high risk of CKD | |
| 28904068 | Wanner (2018) | RCT | EMPA-REG OUTCOME | 4687 | 2333 | Kidney function biomarkers | None | |
| 33084149 | Inzucchi (2021) | RCT | EMPA-REG MET | 430 | 207 | Adiposity markers; Glycaemic markers; | Higher HbA1c at baseline was | |

| | | | | | | | |
|---|---|---|---|---|---|---|---|
| | | | | | | Systolic Blood Pressure | significantly associated with greater HbA1c reduction |

Supplementary 1f. GLP1RA and glycaemic outcomes

| PMID | Author (year) | Study type | Data Source | Intervention (n) | Comparator (n) | Biomarkers examined | Significant interactions | Notes |
|---|---|---|---|---|---|---|---|---|
| 33452595 | Yang 2021 | RCT | ChiCTR-IPR-15006558 | 190 | 0 | Serum FGF21 | High baseline FGF21 levels are associated with poor glycaemic response to exenatide in patients with type 2 diabetes. | |
| 27027802 | Wolffenbuttel 2016 | RCT | NCT00960661; NCT00765817 | 452 | 434 | Adiposity markers | None | |
| 35311356 | Geng 2022 | RCT | CONFIDENCE | 100 | 0 | Genetics | SNP rs163184 in the gene *KCNQ1* was associated with reduced glycaemic response to exenatide in T2DM patients. | |
| 25619391 | Feng 2015 | RCT | NA | 328 | 0 | Age; Adiposity biomarkers; Glycaemic biomarkers; Kidney function biomarkers; Fasting c peptide; Fasting insulin | Patients with high insulin-secreting ability, hyperglucagonemia, and short-duration diabetes may obtain better glycaemic control with liraglutide. | |
| 27161178 | Boustani 2016 | RCT | AWARD | 3136 | 2035 | Age | None | |
| 29573139 | Frias 2018 | RCT | DURATION 8 | 461 | 233 | Age; Sex; Diabetes duration; Ethnicity; Adiposity biomarkers; Glycaemic biomarkers; Kidney function biomarkers | Treatment-by-subgroup interaction was observed for HbA1c change by baseline age subgroup, and HbA1c reductions were greater for patients with higher eGFR. Baseline BMI, T2D duration, sex, race, and ethnicity did not affect HbA1c reductions. | Small group sizes for >65 years and race make those results difficult to conclusively interpret. |
| 32277401 | Yu 2020 | Meta-analysis | AWARD-CHN1; AWARD-CHN2 | 766 | 381 | Glycaemic biomarkers | Greater HbA1c reductions in patients with a higher baseline HbA1c | |
| 35112504 | Yabe 2022 | Meta-analysis | PIONEER 9; PIONEER 10 | 652 | 49 | Adiposity biomarkers; Glycaemic biomarkers; Background anti-hyperglycaemic therapy | HbA1c reductions increased as baseline HbA1c increased. There were no other patterns between the variables investigated and HbA1c changes. | |
| 27265893 | Wysham 2016 | Meta-analysis | AWARD 1 - 6 | 2806 | 0 | Age; Sex; Diabetes duration; Ethnicity; Adiposity biomarkers; Glycaemic biomarkers; Kidney function | Higher baseline HbA1c was associated with greater HbA1c reduction. Age ≤65 years, lower FSG level, FSI level ≤55 pmol/L and eGFR ≤100 mL/min/1.73 | |

| PMID | Author (year) | Study type | Data Source | Intervention (n) | Comparator (n) | Biomarkers examined | Significant interactions | Notes |
|---|---|---|---|---|---|---|---|---|
| | | | | | | biomarkers; History of CVD; Fasting C peptide; Fasting serum insulin | m$^2$ were associated with greater decreases in HbA1c, but the effects were very small. | |
| 33404200 | Terauchi 2020 | Meta-analysis | LixiLan JP-O1; LixiLan JP-O2; LixiLan JP-L | 835 | 516 | Age; Adiposity biomarkers; Glycaemic biomarkers; Background anti-hyperglycaemic therapy | None | |
| 27412701 | Shomali 2017 | Meta-analysis | LEAD 1 - 6; 1860-LIRA-DPP-4 | 2698 | 524 | Ethnicity | None | |
| 28573708 | Shaw 2017 | Meta-analysis | HEELA; EUREXA; NCT00577824; NCT00082381; Nauck 2006; Moretto 2008; NCT00434954; Apovian 2010; DURATION 2; Davies 2012; DURATION-3; DURATION-4; NCT00917267 | 2355 | 0 | Age; Sex; Diabetes duration; Ethnicity; Adiposity biomarkers; Glycaemic biomarkers; Kidney function biomarkers; Background anti-hyperglycaemic therapy | Baseline HbA1c correlated with change in HbA1c. Asian ethnicity and older age were also significantly associated with high glycaemic response to exenatide twice daily. | |
| 26679282 | Seufert 2016 | Meta-analysis | LEAD 1-6; Lira-DPP-4i | 2689 | 519 | Diabetes duration | None | |
| 29748996 | Petri 2018 | Meta-analysis | SUSTAIN 1-3; SUSTAIN-Japan OAD combination | 1935 | 1048 | Adiposity biomarkers; Glycaemic biomarkers; Sex | Greater effects of semaglutide on HbA1c in the 10% of participants with the lowest body weight than in those with the 10% highest weight. | |
| 26662611 | Montanya 2015 | Meta-analysis | LEAD 1-6; Lira-DPP-4i | 2698 | 524 | Adiposity biomarkers | A modest, clinically non-relevant, association between baseline BMI and HbA1c reduction (10kg/m$^2$ increase in baseline BMI corresponds with a 1.1mmol/mol greater HbA1c reduction). | |
| 29603872 | Mathieu 2018 | Meta-analysis | AWARD 1; AWARD 3; AWARD 6 | 817 | 0 | HOMA2-%B (low, middle and high tertiles) | The low tertile of HOMA2-%B coring individuals experienced larger reductions in HbA1c compared to the high tertile when treated with dulaglutide, however this effect was removed when baseline HbA1c was included as a covariate. Greater decreases in fasting blood glucose and | |

| PMID | Author (year) | Study type | Data Source | Intervention (n) | Comparator (n) | Biomarkers examined | Significant interactions | Notes |
|---|---|---|---|---|---|---|---|---|
| | | | | | | | greater increases in fasting C-peptide were observed in the lowest HOMA2-%B tertile. | |
| 22193143 | Henry 2011 | Meta-analysis | NA | NA | NA | Glycaemic biomarkers | Reductions in HbA1C levels and HbA1c goal attainment were greater in groups with higher baseline A1C values. | |
| 31055780 | Gentilella 2019 | Meta-analysis | AWARD 1; AWARD 5; AWARD 6 | 2040 | 315 | Glycaemic biomarkers | Greater HbA1c reductions in patients with higher baseline HbA1c values. | |
| 28817231 | Gallwitz 2017 | Meta-analysis | AWARD 1-6; AWARD 8 | 3375 | 0 | Sex; Diabetes duration; Glycaemic Biomarkers | Greater HbA1c and FBG reductions in patients with a higher baseline HbA1c. | |
| 26594250 | Eto 2015 | Meta-analysis | GetGoal-Duo1; GetGoal-L; GetGoal-L-Asia | 662 | 0 | Adiposity biomarkers | T2D patients in the lowest BMI group relative to those in the highest BMI group had a smaller reduction in HbA1c. | |
| 31769496 | DeSouza 2020 | Meta-analysis | SUSTAIN 1-5; SUSTAIN 7 | 3074 | 0 | Ethnicity | None | |
| 26936426 | Davidson 2016 | Meta-analysis | LEAD 3; LEAD 4; LEAD 6; 1860-LIRA-DPP-4 | 1755 | 642 | Ethnicity | None | |
| 21700561 | Davidson 2011 | Meta-analysis | LEAD 1-6 | NA | NA | Kidney function biomarkers | None | |
| 28303626 | Bonadonna 2017 | Meta-analysis | GetGoal-M; GetGoal-P; GetGoal-S | 546 | 0 | HOMA-B Index (Low vs High) | None | |
| 27767249 | Blonde 2017 | Meta-analysis | GetGoal-M; GetGoal-L; GetGoal-Mono; GetGoal-S; GetGoal-P; GetGoal-Duo 1; GetGoal-X; GetGoal-F1; GetGoal-M-Asia; GetGoal-L-Asia | 2493 | 1465 | Age; Sex; Diabetes duration; Adiposity biomarkers; Glycaemic biomarkers; Background anti-hyperglycaemic therapy | Higher baseline HbA1 was predictive of increased HbA1c reduction. | |
| 35330424 | Kyriakidou 2022 | Observational | Medical records | 116 | 0 | Age; Sex; Diabetes duration; Adiposity biomarkers; Glycaemic biomarkers; Background anti-hyperglycaemic therapy; Genetics | Higher baseline HbA1c and lower baseline weight were associated with better glycaemic response to liraglutide. | CTRB1/2 rs7202877 polymorphism not associated with differential glycaemic response. |
| 29076038 | Berkovic 2017 | Observational | Six tertiary and secondary hospital centres in Croatia | 207 | 0 | Sex; Glycaemic biomarkers; Cardiovascular biomarkers; Background anti- | Independent predictors of durability of HbA1c reduction were initial BMI, HbA1c, systolic BP, and | |

| PMID | Author (year) | Study type | Data Source | Intervention (n) | Comparator (n) | Biomarkers examined | Significant interactions | Notes |
|---|---|---|---|---|---|---|---|---|
| | | | | | | hyperglycaemic therapy | cholesterol. Female gender and shorter duration of diabetes were independent predictors of greater HbA1c reduction. | |
| 31240562 | Yoo 2019 | Observational | Asan Medical Center, Republic of Korea | 234 | 0 | Age; Sex; Diabetes duration; Adiposity biomarkers; Glycaemic biomarkers; Kidney function biomarkers; Dyslipidaemia; Fasting C peptide | Baseline HbA1c was a significant predictor of glycaemic response to dulaglutide. | |
| 30883264 | Yu 2019 | Observational | NA | 285 | 0 | Genetics | The variant allele T of rs10305420 within the GLPR gene was associate with a 0.4% smaller HbA1c reduction after 6 month of exenatide treatment | |
| 35383100 | Yale 2022 | Observational | SURE Canada; SURE Denmark/Sweden; SURE Switzerland; SURE UK | 1212 | 0 | Age; Diabetes duration; Adiposity biomarkers; Glycaemic biomarkers; Previous GLP1RA use | Larger reductions of HbA1c in GLP-1RA-naïve versus GLP-1RA switchers; larger reductions in HbA1c for patients with higher versus lower baseline HbA1c. | |
| 32176827 | Wang 2020 | Observational | Hospital of Xuzhou Medical University, Xuzhou, China | 148 | 0 | Age; Sex; Diabetes duration; Adiposity biomarkers; Glycaemic biomarkers; Background anti-hyperglycaemic therapy; Smoking status; Alcohol intake; Family history of diabetes; Fasting serum insulin; Postprandial serum insulin; HOMA-IR; HOMA-B | Baseline HbA1C and duration of diabetes were identified as predictors of HbA1C reduction. | |
| NA | Thong 2015 | Observational | Association of British Clinical Diabetologists Nationwide Liraglutide Audit | 937 | 0 | Diabetes duration; Background anti-hyperglycaemic therapy | Insulin use and longer diabetes duration, but not the number of OADs taken, predicted a smaller glycaemic response to liraglutide. | |

| PMID | Author (year) | Study type | Data Source | Intervention (n) | Comparator (n) | Biomarkers examined | Significant interactions | Notes |
|---|---|---|---|---|---|---|---|---|
| 24246138 | Thong 2014 | Observational | Specialist diabetes centres in the UK | 116 | 0 | Age; Sex; Diabetes duration; Ethnicity; Adiposity biomarkers; Glycaemic biomarkers; Kidney function biomarkers; Background anti-hyperglycaemic therapy; Urinary C peptide creatinine ratio | Postprandial urinary C-peptide creatinine ratios before and during liraglutide treatment were weakly associated with the glycaemic response to treatment. | |
| 29527621 | Simioni 2018 | Observational | ReaL | 1325 | 0 | Age; Sex; Diabetes duration; Adiposity biomarkers; Glycaemic biomarkers; Kidney function biomarkers; Background anti-hyperglycaemic therapy; Presence of diabetes complications; Hypertension; Dyslipidaemia | Higher baseline HbA1c and shorter T2D duration were predictive of better glycaemic response to liraglutide. | |
| 28983857 | Nunes 2017 | Observational | Optum's electronic health records database | 5361 | 0 | Ethnicity | None | |
| 27889301 | McAdam-Marx 2016 | Observational | National electronic medical record data | 5141 | 0 | Glycaemic biomarkers; Background anti-hyperglycaemic therapy | Greater HbA1c reductions occurred in insulin-naive patients with baseline HbA1c ≥7.0%. | |
| 31920354 | Lee 2019 | Observational | NA | 120 | 0 | Age; Sex; Diabetes duration; Adiposity biomarkers; Glycaemic biomarkers; Kidney function biomarkers; Cardiovascular biomarkers; History of CVD; Fasting C peptide; ALT; AST | Higher baseline HbA1c was associated with a greater reduction in HbA1c. | |
| 25626486 | Lapolla 2015 | Observational | Outpatient units in Veneto Region, Italy | 481 | 0 | Age; Sex; Diabetes duration; Adiposity biomarkers; Cardiovascular biomarkers; Background anti- | HbA1c reduction did not differ among baseline BMI classes. HbA1c <7% goal attainment was predicted by previous metformin | |

| PMID | Author (year) | Study type | Data Source | Intervention (n) | Comparator (n) | Biomarkers examined | Significant interactions | Notes |
|---|---|---|---|---|---|---|---|---|
| | | | | | | hyperglycaemic therapy; Glycaemic biomarkers; Previous anti-hyperglycaemic drug use | monotherapy and insulin naivety. | |
| 29341370 | Gorgojo-Marti-nez 2017 | Observational | Four tertiary Spanish hospitals | 148 | 0 | Age; Sex; Diabetes duration; Adiposity biomarkers; Glycaemic biomarkers; Kidney function biomarkers; Cardiovascular biomarkers; History of CVD; Background anti-hyperglycaemic therapy; Microvascular complications; Smoking status; AST; ALT; Background antihypertensive and lipid-lowering drug use. | Higher HbA1c was associated with increased glycaemic response to exenatide. | |
| 29943854 | Gomez-Peralta 2018 | Observational | Six Spanish centres | 799 | 0 | Age; Sex; Diabetes duration; Adiposity biomarkers; Glycaemic biomarkers; Cardiovascular biomarkers; Background anti-hyperglycaemic therapy | Longer treatment with liraglutide was a predictor of improved HbA1c response, whereas higher baseline HbA1c, longer Type 2 diabetes duration and treatment with insulin were predictors of worse HbA1c response. | |
| 26830854 | Gimeno-Orna 2016 | Observational | NA | 117 | 0 | ALT | Elevated baseline transaminase values and decreased transaminase levels during follow-up are associated to a favourable glycaemic response to GLP-1 RAs | |
| 32753164 | Dalmazi 2020 | Observational | An outpatient diabetes clinic in Italy | 186 | 0 | Age; Sex; Diabetes duration; Adiposity biomarkers; Glycaemic biomarkers; Kidney function biomarkers; Cardiovascular biomarkers; Background | Predictors of adequate glycaemic response were shorter diabetes duration and not switching to a different GLP-1RA, respectively | |

| PMID | Author (year) | Study type | Data Source | Intervention (n) | Comparator (n) | Biomarkers examined | Significant interactions | Notes |
|---|---|---|---|---|---|---|---|---|
| | | | | | | anti-hyperglycaemic therapy | | |
| 29386249 | Dennis 2018 | Observational | PRIBA; CPRD | 4803 | 0 | Adiposity biomarkers; Cardiovascular biomarker; Markers of insulin resistance | No evidence of an association between any marker of insulin resistance and 6-month glycaemic response to GLP-1 receptor agonists; no evidence for a difference in response to GLP-1 receptor agonists across the obesity and triglyceride defined subgroups. | |
| 25245811 | Chitnis 2014 | Observational | General Electric Centricity electronic medical records database | 3005 | 0 | Adiposity biomarkers | None | |
| 29241884 | Brekke 2017 | Observational | Independent administrative claims databases associated with Optimum and HealthCore in the United States | 3773 | 0 | Age; Sex; Adiposity biomarkers; Glycaemic biomarkers; Background anti-hyperglycaemic therapy; Cardiovascular Biomarkers; Geographic location; Quan-modified Charlson comorbidity index score; Specialty of prescribing physician; Diabetes-related hospitalisation; Number of ambulance visits; Neuropathy; Retinopathy; Mental illness; Polypharmacy; Hypoglycaemic event; Improving weight control at baseline. | Polypharmacy and hypoglycaemia were associated with greater HbA1c reductions in response to liraglutide. Individuals 18 to 39 years and those with $HbA_{1c}$ of 7.0% to less than 8.0% had higher persistence with liraglutide. | |
| 32574827 | Berra 2020 | Observational | 5 diabetes centres of the Milan (Italy) | 626 | 0 | Age; Diabetes duration; Glycaemic biomarkers; Pre-study medication | Predictors of the achievement of HbA1c≤7.0 % were low baseline HbA1c and short duration of diabetes. Neither sex nor age had significant effects on any clinical or laboratory outcome. | |

| PMID | Author (year) | Study type | Data Source | Intervention (n) | Comparator (n) | Biomarkers examined | Significant interactions | Notes |
|---|---|---|---|---|---|---|---|---|
| 23630427 | Anichini 2013 | Observational | 5 diabetes outpatient clinics in Tuscany, Italy | 315 | 0 | Sex; Diabetes duration; Adiposity biomarkers; Glycaemic biomarkers; Background anti-hyperglycaemic therapy; B-cell function biomarkers | One-year glycaemic target response was associated with high baseline HbA1c levels and longer diabetes duration among males; concomitant metformin therapy was a predictor of better glycaemic targets among females. | |
| 24877253 | Carrington 2014 | Observational | Medical records in a specialist diabetes clinic | 446 | 0 | Diabetes duration; Glycaemic biomarkers; Background anti-hyperglycaemic therapy; GLP1RA treatment length | Higher baseline HbA1c, longer duration of diabetes, longer exenatide use, and use of insulin or sulfonylureas at study end predicted better glycaemic response. | |
| 33801192 | Mirabelli 2021 | Observational | Unit of Endocrinology and Diabetes at Hospital "Pugliese-Ciaccio" in Catanzaro, Italy | 126 | 0 | Glycaemic biomarkers | Higher baseline HbA1c was a predictor of HbA1c reduction ≥ 0.5%. | |

### Supplementary table 2: Individual Trials included in meta-analyses

#### a: List of individual CVOT and renal outcome trials for SGLT2i included in meta-analysis studies

| Trial name | Main paper PMID | ClinicalTrials.gov | Population | Interventions | Primary outcome | N | % with T2D | Median fu, years |
|---|---|---|---|---|---|---|---|---|
| EMPA-REG | 26378978 | NCT01131676 | Established CVD | Empagliflozin 25mg vs. 10mg vs. placebo | 3-point MACE | 7,020 | 100% | 3.1 |
| CANVAS* | 28605608 | NCT01032629 | Established CVD or high CVD risk | Canagliflozin 300mg vs. 100mg vs. placebo | 3-point MACE | 4,330 | 100% | 5.7 |
| CANVAS-R* | 28605608 | NCT01989754 | Established CVD or high CVD risk | Canagliflozin 300mg vs. 100mg vs. placebo | Progression of albuminuria | 5,812 | 100% | 2.1 |
| DECLARE | 30415602 | NCT01730534 | Established CVD or high CVD risk | Dapagliflozin 10 mg vs. placebo | 3-point MACE | 17,160 | 100% | 4.2 |
| CREDENCE | 30990260 | NCT02065791 | eGFR 30-90 and albuminuria | Canagliflozin 100mg vs. placebo | Composite renal | 4,401 | 100% | 2.6 |
| VERTIS-CV | 32966714 | NCT01986881 | Established CVD | Ertugliflozin 15mg vs. 5mg vs. placebo | 3-point MACE | 8,246 | 100% | 3.5 |
| DAPA-HF | 31535829 | NCT03036124 | Established HF | Dapagliflozin 10 mg vs. placebo | HF composite #1 | 4,744 | 42% | 1.5 |
| SCORED | 33200891 | NCT03315143 | eGFR 25-60 | Sotagliflozin 200-400mg vs. placebo | HF composite #1 | 10,584 | 100% | 1.3 |
| EMPEROR-P | 34449189 | NCT03057951 | Established HF | Empagliflozin 10mg vs. placebo | HF composite #2 | 5,988 | 49% | 2.2 |
| SOLOIST-WHF | 33200892 | NCT03521934 | Recent HF hospitalization | Sotagliflozin 200-400mg vs. placebo | HF composite #3 | 1,222 | 100% | 0.8 |
| EMPEROR-R | 32865377 | NCT03057977 | Established HF | Empagliflozin 10mg vs. placebo | HF composite #2 | 3,730 | 50% | 1.3 |
| DAPA-CKD | 32970396 | NCT03036150 | eGFR 25-75 | Dapagliflozin 10 mg vs. placebo | Composite renal | 4,304 | 68% | 0.75 |

3-point MACE: cardiovascular death, non-fatal MI, non-fatal stroke
HF composite #1: hospitalization or an urgent visit with intravenous therapy for heart failure or cardiovascular death
HF composite #2: hospitalization for heart failure or cardiovascular death
HF composite #3: hospitalization or an urgent visit for heart failure or cardiovascular death
*Typically analyzed together

#### b: Summary of CVOTs for GLP1RA included in meta-analysis studies

| Trial name | Main paper PMID | ClinicalTrials.gov | Population | Interventions | Primary outcome | N | % with T2D | Median fu, years |
|---|---|---|---|---|---|---|---|---|
| ELIXA | 26630143 | NCT01147250 | Established CVD | Lixisenatide 1.8 mg vs. placebo | 4-point MACE | 6,068 | 100% | 2.1 |
| EXSCEL | 28910237 | NCT01144338 | General T2D | Exenatide 2 mg weekly vs. placebo | 3-point MACE | 14,752 | 100% | 3.2 |
| HARMONY | 30291013 | NCT02465515 | Established CVD | Albiglutide 30–50 mg weekly vs placebo | 3-point MACE | 9463 | 100% | 1.6 |
| LEADER | 27295427 | NCT01179048 | Established CVD or high CVD risk | Liraglutide 1.8 mg vs.placebo | 3-point MACE | 9,340 | 100% | 3.8 |
| PIONEER-6 | 31185157 | NCT02692716 | Established CVD or high CVD risk | Semaglutide 14mg daily vs placebo | 3-point MACE | 3,183 | 100% | 1.3 |
| SUSTAIN 6 | 28249135 | NCT01720446 | Established CVD/HF/CKD or high CVD risk | Semaglutide 0.5 mg vs. 1.0 mg vs. placebo | 3-point MACE | 3,297 | 100% | 2.1 |
| REWIND | 31189511 | NCT01394952 | Established CVD or high CVD risk | Dulaglutide 1.5 mg weekly vs. placebo | 3-point MACE | 9,901 | 100% | 5.4 |

3-point MACE: cardiovascular death, non-fatal MI, non-fatal stroke
4-point MACE: as above plus hospitalisation for unstable angina

#### Supplementary Figure 2 – Heat Map of quality assessment for included studies

##### Supplementary Figure 2a

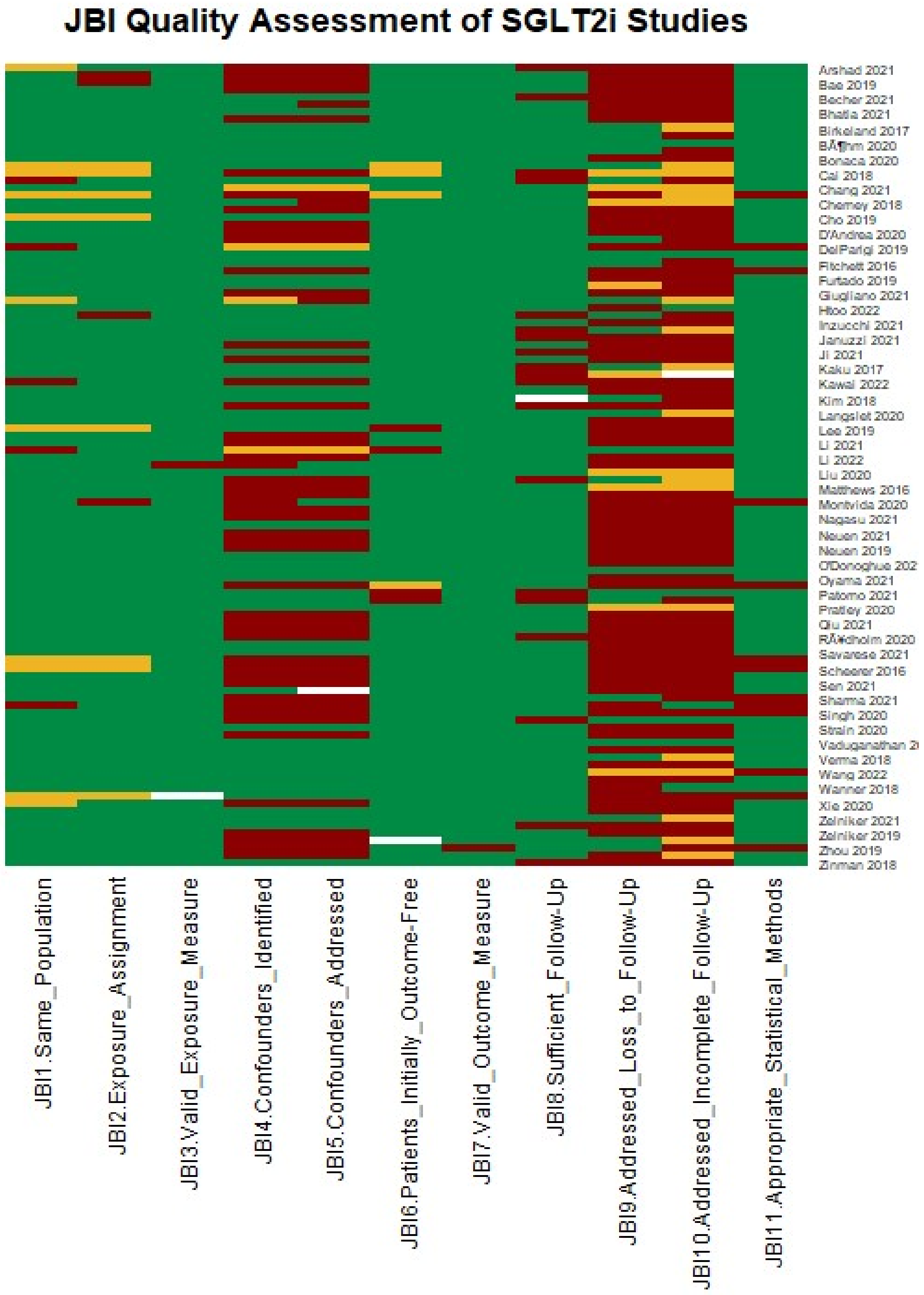

#### Supplementary Figure 2b

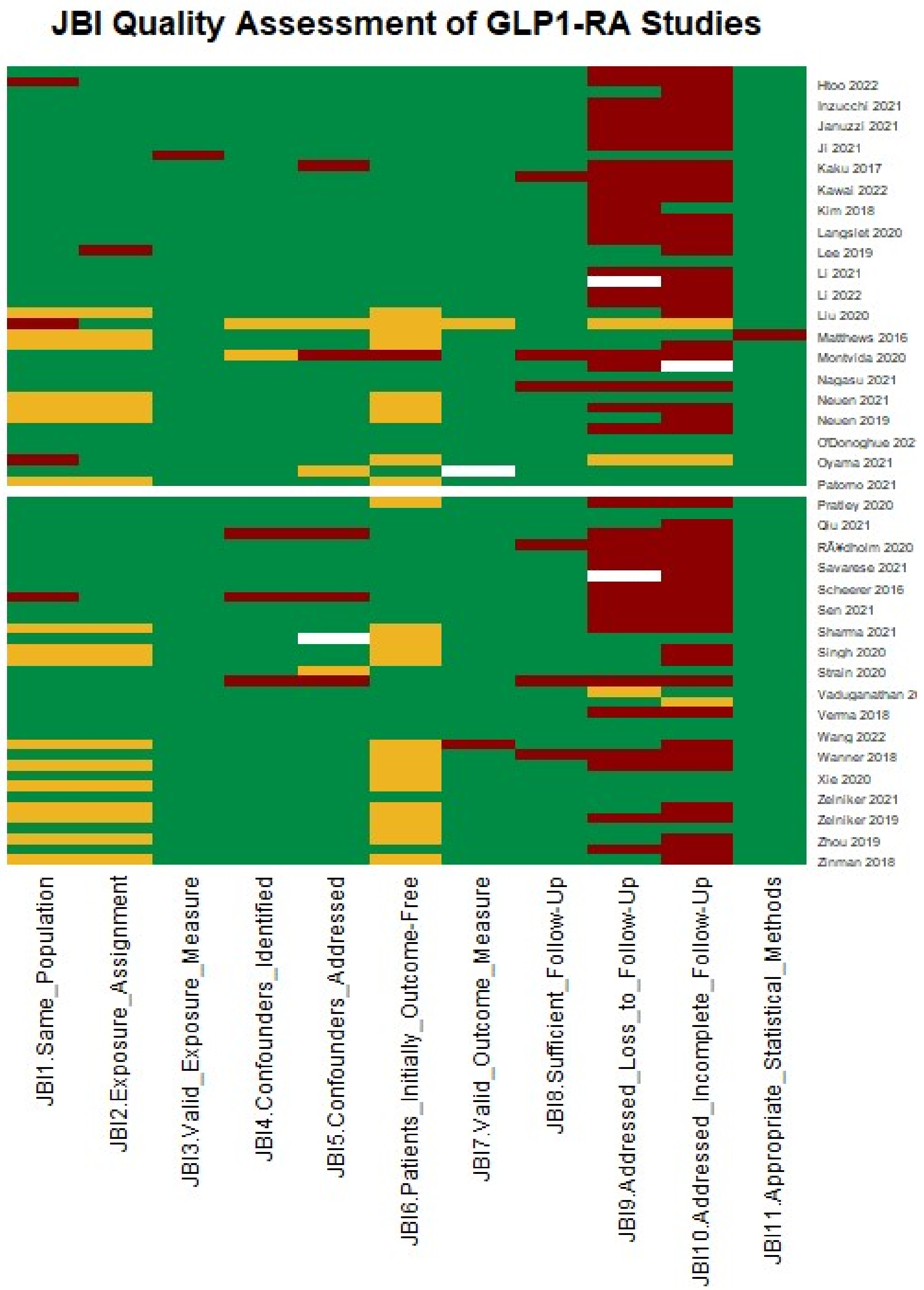
